# Virological and serological characterization of critically ill patients with COVID-19 in the UK: a special focus on variant detection

**DOI:** 10.1101/2021.02.24.21251989

**Authors:** Jeremy Ratcliff, Dung Nguyen, Matthew Fish, Jennifer Rhynne, Aislinn Jennings, Sarah Williams, Farah Al-Beidh, David Bonsall, Amy Evans, Tanya Golubchik, Anthony C Gordon, Abigail Lamikanra, Pat Tsang, Nick Ciccone, Ullrich Leuscher, Wendy Slack, Emma Laing, Paul R Mouncey, Sheba Ziyenge, Marta Olivera, Rutger Ploeg, Kathryn M Rowan, Manu Shankar-Hari, David J. Roberts, David K Menon, Lise Estcourt, Peter Simmonds, Heli Harvala on behalf of the REMAP-CAP Immunoglobulin Domain UK Investigators (listed in the Supp. Appendix).

**Author notes:** shared last authors (Peter Simmonds, Heli Harvala).

## Abstract

**Background:** Treatment of COVID-19 patients with convalescent plasma containing neutralising antibody to SARS-CoV-2 is under investigation as a means of reducing viral loads, ameliorating disease outcomes, and reducing mortality. However, its efficacy might be reduced in those infected with the emerging B.1.1.7 SARS-CoV-2 variant. Here, we report the diverse virological characteristics of UK patients enrolled in the Immunoglobulin Domain of the REMAP-CAP randomised controlled trial.

**Methods:** SARS-CoV-2 viral RNA was detected and quantified by real-time PCR in nasopharyngeal swabs obtained from study subjects within 48 hours of admission to intensive care unit. Antibody status was determined by spike-protein ELISA. B.1.1.7 strain was differentiated from other SARS-CoV-2 strains by two novel typing methods detecting the B.1.1.7-associated D1118H mutation with allele-specific probes and by restriction site polymorphism (SfcI).

**Findings:** Of 1260 subjects, 90% were PCR-positive with viral loads in nasopharyngeal swabs ranging from 72 international units [IUs]/ml to 1.7×10^11^ IU/ml. Median viral loads were 45-fold higher in those who were seronegative for IgG antibodies (n=314; 28%) compared to seropositives (n=804; 72%), reflecting in part the latter group’s possible later disease stage on enrolment. Frequencies of B.1.1.7 infection increased from early November (<1%) to December 2020 (>60%). Anti-SARS-CoV-2 seronegative individuals infected with wild-type SARS-CoV-2 had significantly higher viral loads than seropositives (medians of 1.2×10^6^ and 3.4 ×10^4^ IU/ml respectively; p=2×10^−9^). However, viral load distributions were elevated in both seropositive and seronegative subjects infected with B.1.1.7 (13.4×10^6^ and 7.6×10^6^ IU/ml; p=0.18).

**Interpretation:** High viral loads in seropositive B.1.1.7-infected subjects are consistent with increased replication capacity and/or less effective clearance by innate or adaptive immune response of B.1.1.7 strain than wild-type. As viral genotype was associated with diverse virological and immunological phenotypes, metrics of viral load, antibody status and infecting strain should be used to define subgroups for analysis of treatment efficacy.

## INTRODUCTION

The catastrophic zoonotic emergence of severe acute respiratory syndrome coronavirus 2 (SARS-CoV-2) in the human population in China at the end of 2019, and its subsequent pandemic spread has caused global devastation. By the end of January 2021, over 100 million infections and over 2 million COVID-19-associated deaths have been reported worldwide To date, only limited treatment regimens exist for COVID-19 patients^1,2^, and their management is primarily supportive, with case-fatality rates remaining at 1% - 2% in Western countries including the UK^3^. In the search of alternative methods to treat COVID-19, convalescent plasma therapy boosting levels of neutralising antibody has been considered as a potential means to reduce morbidity and mortality^4,5^, particularly if given in early stages of infection^6^. RECOVERY and REMAP-CAP are randomized controlled trials investigating the efficacy of convalescent plasma therapy conducted in the UK with REMAP-CAP also recruiting participants globally^6,7^. In the REMAP-CAP trial^7^ patients with severe COVID-19, restricted to those in intensive care units (ICUs), are transfused with one to two plasma units collected from donors with previous documented SARS-CoV-2 infection and confirmed to have high plasma titres of neutralising antibody (>1:100 based on in vitro live virus neutralisation assays) using surrogate testing with a spike antigen-based ELISA^8^.

While final statistical analysis is awaited, both trials have been paused as an initial analysis did not show a significant benefit of treatment across the overall study population in terms of either COVID-19 associated mortality or number of organ-support free days^9,10^. However, analysis of the data will need to account for patient variables that may influence treatment efficacy, and address patient outcomes other than death. For example, the potential efficacy of convalescent plasma therapy may be influenced by disease stage at trial enrolment. Severe COVID-19 disease may be driven by either the direct effects of virus replication in the respiratory tract, or secondarily from damage associated with the often intense inflammatory antiviral response that usually leads to clearance of SARS-CoV-2. Treatment with neutralising antibody may primarily influence outcomes during the early stages of active virus replication-induced disease but could be less relevant in those patients who have cleared their infection and seroconverted for anti-SARS-CoV-2 antibody. Therefore, we have determined the pre-treatment viral loads in respiratory samples and serological status to categorise study subjects by their disease stage.

Another variable influencing treatment outcomes may originate from effects of strain variation of SARS-CoV-2. A variant of SARS-CoV-2 with a D614G mutation in the spike protein has been described^11,12^ along with more recent mutants such as B.1.1.7 in the UK, B.1.351 in South Africa and B.1.1.28 in Brazil^13^, all potentially more transmissible^14^; furthermore, B.1.1.7 may result in a higher mortality than the original pandemic strain^15^. As regards B.1.351 (South Africa), clustered amino acid changes in the spike protein gene may render this mutant partially antigenically distinct from the prototype strain^16^. These virological differences, including the possibility of antigenic escape from neutralising antibodies in convalescent plasma collected from individuals with earlier strains of SARS-CoV-2, may individually or collectively affect the efficacy of convalescent plasma therapy. To identify SARS-CoV-2 strains, we developed a real-time PCR targeting the D1118H polymorphism in the spike gene that is characteristic of the B.1.1.7 strain. This provided a rapid means to differentiate B.1.1.7 from other SARS-CoV-2 strains; in addition, the assay was found to be more effective than high throughput sequencing for low viral load samples. A simpler agarose gel-based method was also developed and evaluated to enable B.1.1.7 identification in resource-limited settings.

Using these methods, we report substantial variability in pre-treatment viral loads, variability in serological status and an increasing detection of B.1.1.7 during the enrolment period of UK participants in the international REMAP-CAP trial^7^. Measurement of these variables will be of considerable value in analysing effects of convalescent plasma therapy and potentially identifying sub-groups of patients for whom this treatment may be effective.

## MATERIALS AND METHODS

### Patient Recruitment and Sample Collection

All subjects were enrolled in the UK and had a laboratory confirmed diagnosis of SARS-CoV-2 infection with concomitant severe pneumonia requiring ICU admission. Patients were not eligible if more than 48 hours had elapsed since their ICU admission, if they had already received treatment with any other non-trial prescribed antibody therapy (monoclonal antibody, hyperimmune immunoglobulin, or convalescent plasma) or if more than 14 days had elapsed since hospital admission. A total of 122 hospitals have recruited patients in this trial in the UK (Table S1; Suppl. Data)^17^. Nasopharyngeal or oropharyngeal swabs and serum samples for baseline virological testing were collected from patients prior to the enrolment and frozen at −80°C.

### Ethical statement

The study was conducted according to the principles of the latest version of the Declaration of Helsinki (version Fortaleza 2013), and in accordance with regulatory and legal requirements (EudraCT number: 2015-002340-14). The study was approved by London-Surrey Borders Research Ethics Committee London Centre (18/LO/0660). Written or verbal informed consent, in accordance with regional legislation, is obtained from all patients or their surrogates.

### Nucleic Acid Extraction

Viral RNA was extracted from patient samples using the QIAamp Viral RNA Mini Kit (QIAGEN). 140 µL of respiratory sample was mixed with 560 µL of Buffer AVL containing 20 µg/mL of linear polyacrylamide (ThermoFisher Scientific), extracted according to the manufacturer’s spin protocol, and eluted into 60 µL of buffer AVE. Samples that arrived dry were resuspended in 2 mL of PBS and incubated at room temperature for 20 minutes prior to extraction. NIBSC research reagent 19/304 (https://nibsc.org/products/brm_product_catalogue/detail_page.aspx?catid=20/146) was extracted using the QIAamp Viral RNA Mini Kit and serially diluted in RNA storage solution (Thermo Fisher Scientific; 1⍰mM sodium citrate, pH 6.4) containing herring sperm carrier RNA (50⍰µg/mL) and RNasin (New England BioLabs UK, 100⍰U/mL). All samples were subject to a single freeze-thaw cycle between extraction and RT-PCR quantification.

### Real-time Polymerase Chain Reaction (PCR)

Viral RNA was detected and quantified by real-time polymerase chain reaction (RT-PCR) using the Quantitect Probe RT-PCR kit (QIAGEN) with CDC N1 primers (Table 1; https://www.cdc.gov/coronavirus/2019-ncov/lab/rt-pcr-panel-primer-probes.html) (final concentrations 500 nM for primers and 125 nM for probe (ATDBio)) in 20 µL reaction volumes and 5 µL of extracted RNA. RT-PCR was carried out on an Applied Biosystems StepOnePlus Real-Time PCR system (ThermoFisher Scientific) with the following settings: 50°C for 30m, 95°C for 15m, and 40 cycles of 94°C for 15s and 60°C for 1m. Intra-assay variation was standardized through use of a standard curve of NIBSC RNA control 19/304 serially diluted from 10,000 copies/reaction to 100 copies/reaction. RT-PCR Ct values were converted to copy number/reaction by use of the standard curve and to international units/mL by the conversion rate provided by NIBSC.

**TABLE 1.** SPECIFICITY AND SENSITVITY OF THE EIGHT POLYMORHISMS IN B.1.1.7 FOR STRAIN IDENTIFICATION

SPECIFICITY AND SENSITIVITY OF THE EIGHT POLYMORPHISMS IN B.1.1.7 FOR STRAIN IDENTIFICATION
| Strain | Source | H69/V70 deletion<sup>1</sup> |  | Y144 deletion |  | N501Y |  | A570D |  |
| --- | --- | --- | --- | --- | --- | --- | --- | --- | --- |
|  |  | WT | Del | WT | Del | A | U | C | A |
| B.1.1.7 | UK<sup>2</sup> | 0 | 26,328 | 0 | 26,328 | 0 | 26,328 | 2 | 26,359 |
| B.1.1.28 | All<sup>3</sup> | 374 | 0 | 374 | 0 | 294 | 80 | 374 | 0 |
| B.1.351 | All<sup>4</sup> | 834 | 0 | 834 | 0 | 0 | 834 | 833 | 0 |
| WT | UK<sup>4</sup> | 36,577 | 470 | 35,708 | 46 | 36,868 | 177 | 37,597 | 0 |
| **Specificity/Sensitivity<sup>5</sup>** |  | 98.73% / 100% |  | 99.87% / 100% |  | 99.52% / 100% |  | 99.98% / 99.99% |  |

| Strain | Source | P681H |  | T716I |  | S982A |  | D1118H |  |
| --- | --- | --- | --- | --- | --- | --- | --- | --- | --- |
|  |  | C | A | C | U | U | G | G | C |
| B.1.1.7 | UK | 0 | 26,351 | 2 | 26,351 | 0 | 26,360 | 0 | 26,358 |
| B.1.1.28 | All | 374 | 0 | 374 | 0 | 374 | 0 | 374 | 0 |
| B.1.351 | All | 832 | 1 | 832 | 1 | 831 | 0 | 834 | 0 |
| WT | UK | 37,442 | 34 | 37,528 | 23 | 37,563 | 0 | 37,579 | 8 |
| **Specificity/Sensitivity** |  | 99.69% / 99.97% |  | 99.94% / 99.99% |  | 100% / 100% |  | 99.98% / 99.97% |  |

<sup>1</sup>For each mutation, mutations associated with B.1.1.7 shown in right hand column
<sup>2</sup>Available sequences from GISAID Nov, Dec, 2020 and Jan, 2021
<sup>3</sup>All available sequence worldwide
<sup>4</sup>Available sequences from GISAID Jan-April, July, Nov, Dec, 2020 and Jan, 2021
<sup>5</sup>Specificity and sensitivity for differentiation B.1.1.7 from WT SARS-CoV-2 sequences

### Typing assays to identify B.1.1.7 SARS-CoV-2 strains

An RT-PCR using allele-specific probes (ASP) and nested PCR Restriction Fragment Length Polymorphism for the G➔C polymorphism at position 24814 (D1118H) (Table S1; Suppl. Data) were used to differentiate wild-type and B.1.1.7 strains. Detailed methods are available as Protocols A and B (Suppl. Data).

### Nucleotide sequencing

Whole virus genomes were sequenced using the virus-specific PCR-free sequencing method veSEQ. The full method description is provided in Protocol C (Suppl. Data).

### Serology

Baseline blood samples were also collected from all patients to determine the presence of SARS-CoV-2 IgG antibodies against spike protein at the time of transfusion. These were determined using the recently developed and validated 386-well plate ELISA coupled with an automated liquid handler^18^.

## RESULTS

### Trial enrolment

Viral loads and anti-SARS-CoV-2 IgG antibody status were determined in respiratory and/or serum samples collected from patients enrolled into REMAP-CAP trial in the UK from 25^th^ May 2020 through 4^th^ January 2021.

### Pre-treatment viral loads

RNA extracted from respiratory samples was amplified by real-time PCR using CDC N1 (nucleoprotein gene) primers (Fig. 1A). Ct values were converted to viral loads through co-amplification and calibration with the SARS-CoV-2 international reference standard (NIBSC, UK). This placed the assay sensitivity threshold at 100 copies/ml for samples with Ct values >35. Using this calibrated assay, a total of 1129 from 1260 samples (89%) were SARS-CoV-2 RNA positive. Viral loads ranged from <100 IU/ml to 1.7 x 10^11^ IU/ml, a remarkable >2 billion-fold range in levels of secreted virus.

**FIGURE 1.**
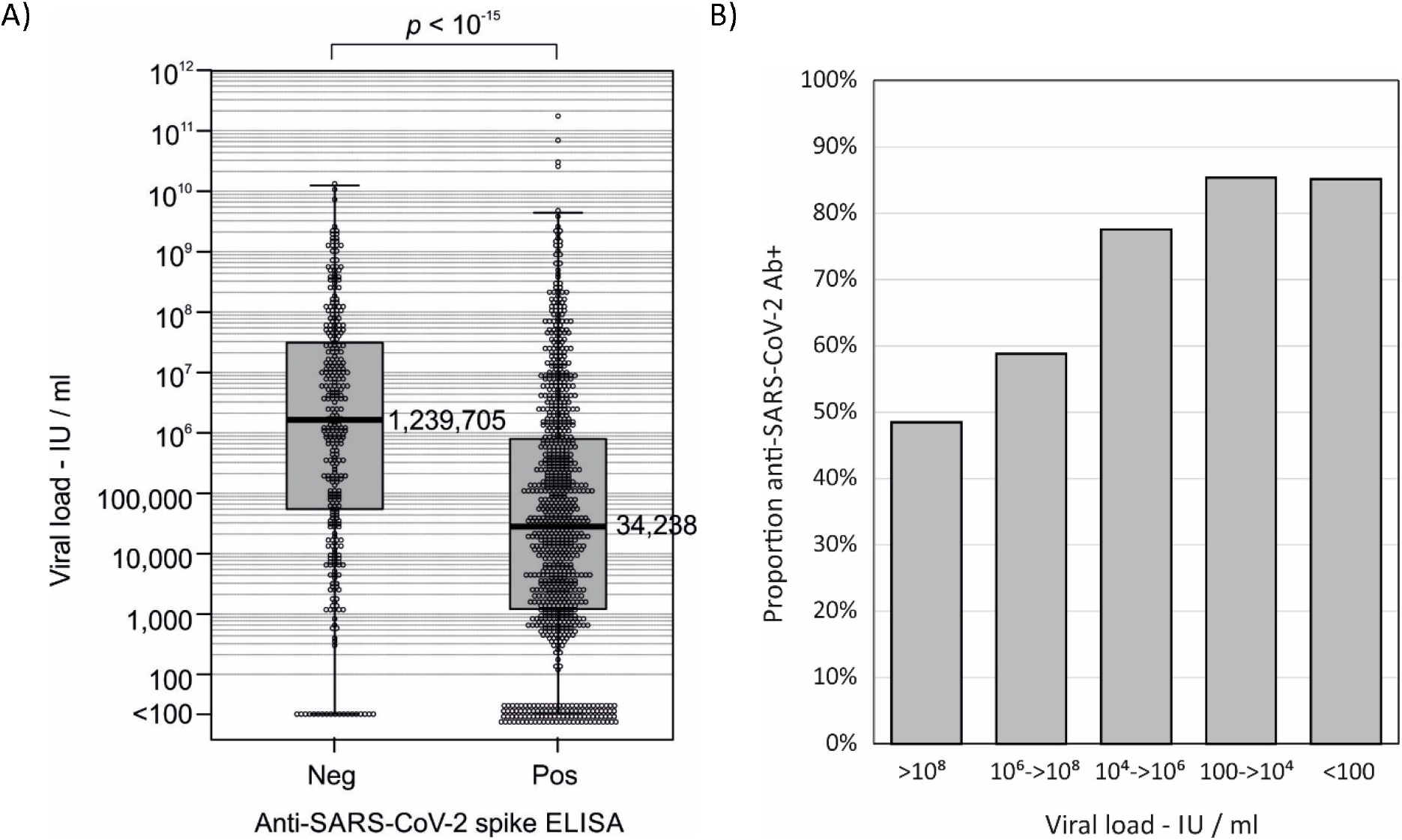
COMPARISON OF VIRAL LOADS IN ANTI-SARS-CoV-2 SEROPOSITIVE AND SERONEGATIVE SUBJECTS (A) Viral loads of seropositive and seronegative study subjects determined by RT-qPCR of pre-treatment respiratory samples. Median values shown to the right of Tukey box plots. Distributions were compared by Kruskall-Wallis non-parametric test. (B) Frequency of seropositivity in individuals with different viral loads quantified in respiratory samples.

### Serology

The serological status of each enrolled patient was determined by ELISA to recombinant spike protein. Results were expressed as positive and negative although the degree of antibody reactivity could not be quantified by the nature of the assay. Antibody status has been determined in 1118 patients included in this study to date, of whom 804 were determined as seropositive (72%). The median viral load of seronegative individuals was 36x higher than seropositives with markedly different distributions of viral loads between the two groups (Fig. 1A). The proportion of seropositive patients varied systematically with viral load ranges, being highest (85%) in those with the lowest viral load (< through to 48% in those with viral loads >10^8^ IU/ml (Fig. 1B).

### SARS-CoV-2 strain identification

As infections with the emerging B.1.1.7 strain may respond differently to treatment compared to previously circulating strains of SARS-CoV-2, two different typing assays were developed to identify B.1.1.7 in trial patients. We first analysed the specificity and predictive value of individual polymorphic sites in the spike gene of B.1.1.7 (H69/V70del., Y144del., N501Y, A570D, P681H, T716I, S982A, D1118H) that differentiated them from other SARS-CoV-2 strains (Table 1). All but one of the defined polymorphisms differentiated B.1.1.7 with very high sensitivity and specificity from previously circulating strains in the UK and from emerging strains in South Africa and Brazil (B.1.351 and B.1.1.28). The exception was the H69/V70 del site which showed increasing frequencies with time in multiple lineages of wild-type strains (Table S2; Suppl. Data). As the G->C change associated with D1118H also led to the loss of a restriction enzyme site (SfcI), amplicons from this region were used in typing assays with allele-specific probes (ASP; Fig. S1A; Suppl. Data) and nested-PCR RFLP assays (Fig. S1B; Suppl. data).

For baseline genomic characterisation, the first 284 respiratory samples collected in November 2020 from the trial patents were sequenced by HTS using SARS-CoV-2-baited target enrichment. Variable coverage of the genome was achieved, associated with viral load (Fig. S2; Suppl. Data). All sequences with coverage over codon 164 of the spike gene showed the D614G mutation. Phylogenetic analysis (Fig. S3; Suppl. Data) of those samples showing >65% sequence coverage demonstrated that the majority fell into the lineage GV (according to its designation in GISAID). Of the sequences obtained, 13 of the 173 sequences with sufficient sequence in the spike gene to enable assignment were identifiable as B.1.1.7. No sequences corresponded to emerging strains B.1.351 (South Africa) or B.1.1.28 (Brazil).

A subset of 40 samples sequenced across the spike gene were used for evaluation of the typing assays. These produced highly concordant results with 100% specificity, 97.5% sensitivity for the ASP assay, and 97.5% specificity, 93.5% sensitivity for the RFLP assay (Table S3; Suppl. Data). Study samples were typed by a combination of the three methods with 521 (400 WT; 121 B.1.1.7) from 645 from tested (81%) successfully typed.

### Emergence and virological associations of SARS-CoV-2 strains

Infections with the B.1.1.7 strain were first detected among enrolled patients in early November and rapidly increased in frequency through November and December, representing over two thirds of infections in the week beginning 21^st^ December 2020 (Fig. 2). Frequencies of B.1.1.7 infections matched their emergence in the general UK population based on analysis of SARS-CoV-2 sequences deposited on GISAID in the corresponding study weeks, albeit at a 1.5-2-fold consistently greater frequency. Geographically, B.1.1.7 infections occurred at greatest frequency in subjects enrolled in South East England (including London) and in East England (approximately 50%; Fig. 3). Cohort enrolment from different regions of England closely matched the proportions of COVID-19 diagnoses reported to PHE over the study period.

**FIGURE 2.**
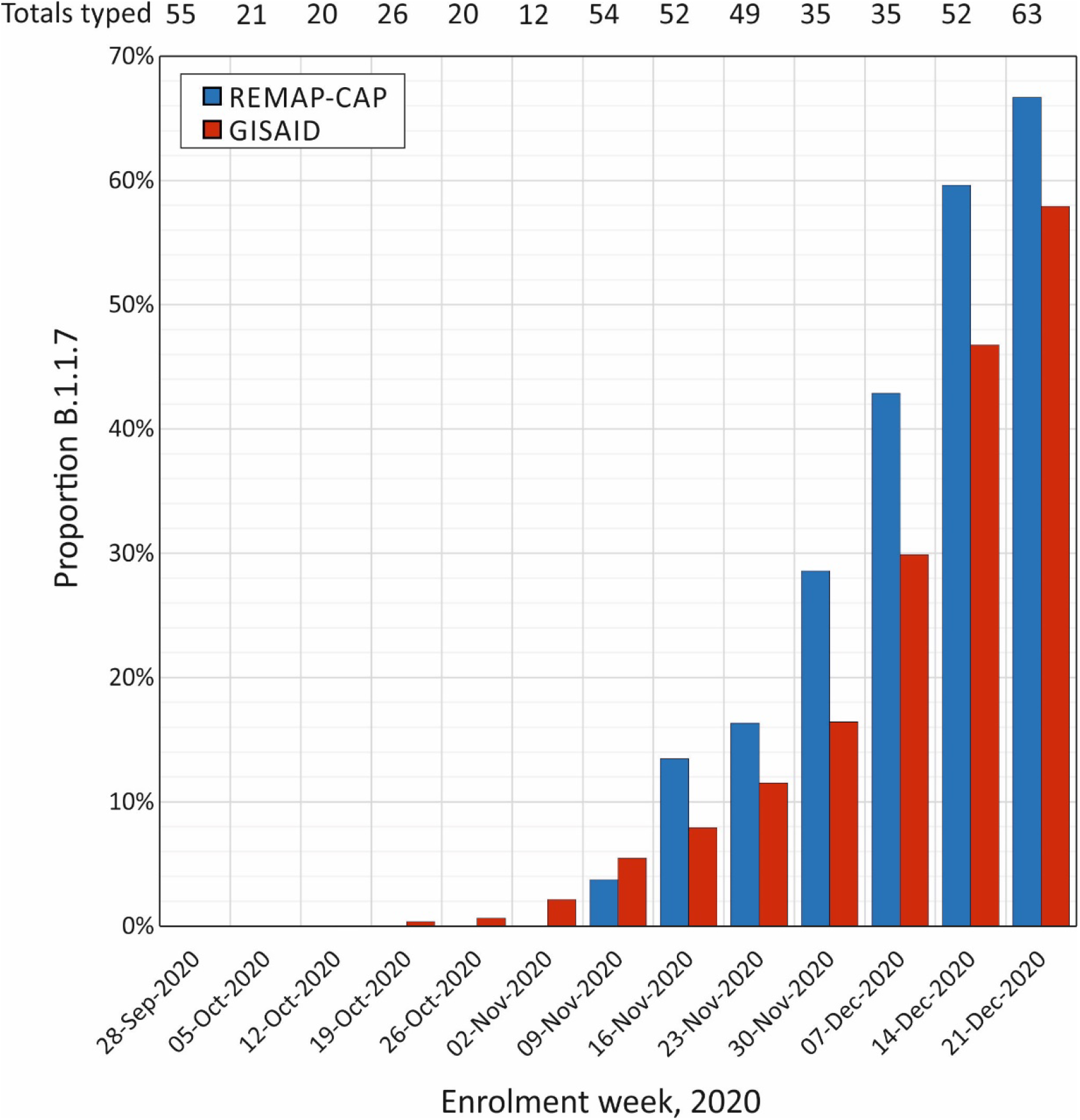
TEMPORAL EMERGENCE OF THE SARS-CoV-2 B.1.1.7 CLADE Proportion of subjects with the B.1.1.7 clade virus enrolled to the REMAP-CAP trial in different weeks over the study period compared to proportions in the wider UK population from sequences deposited in GISAID. Numbers at the top of the graph indicate total enrolments / week.

**FIGURE 3.**
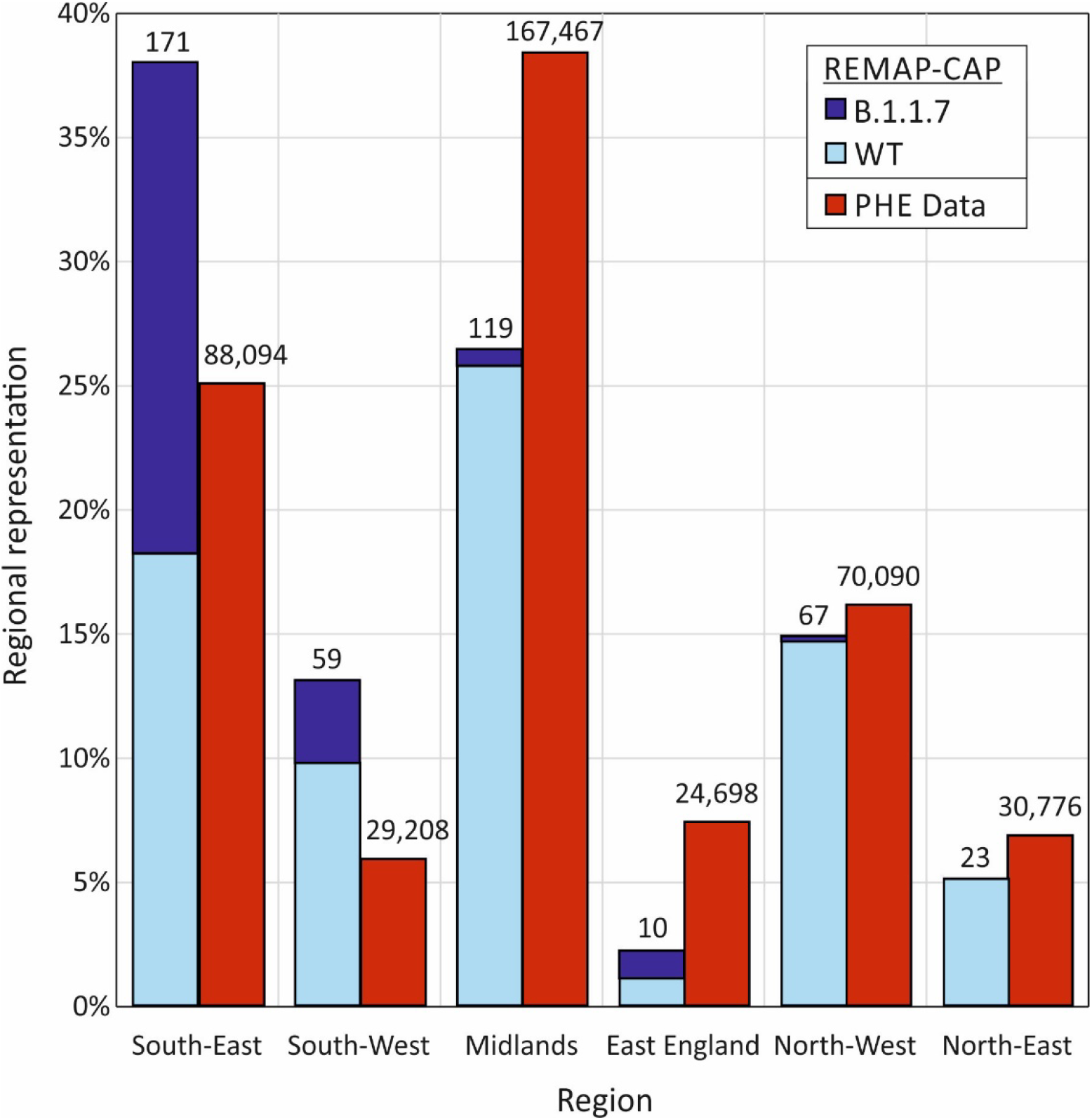
REGIONAL REPRESENTATION OF STUDY SUBJECTS AND SARS-CoV-2 CASES WITH GENOTYPING DATA AVAILABLE IN ENGLAND Regional representation of subjects enrolled into the REMAP-CAP trial compared to distributions of SARS-CoV-2 infections proportions in England regions reported to PHE over the study period. Numbers at the top of the graph indicate enrolments numbers / region.

Viral loads ranges from subjects infected with the B.1.1.7 strain differed substantially from those with wild-type SARS-CoV-2, with medians of 10,146,774 and 820,689 IU/ml respectively (p=2×10^−9^). Remarkably, significant differences in viral loads were only apparent among subjects who were seropositive for SARS-CoV-2 antibody (Fig. 4), with an approximately 40-fold difference between medians of B.1.1.7 and wild-type infections. Contrastingly, viral loads varied by less than 2-fold between those with B.1.1.7 and wild-type infections in seronegative subjects. Seropositivity had no significant effect on B.1.1.7 viral loads (p=0.47). Viral load distributions were elevated in both seropositive and seronegative subjects infected with B.1.1.7 (13.4×10^6^ and 7.6×10^6^ IU/ml; p=0.18).

**FIGURE 4.**
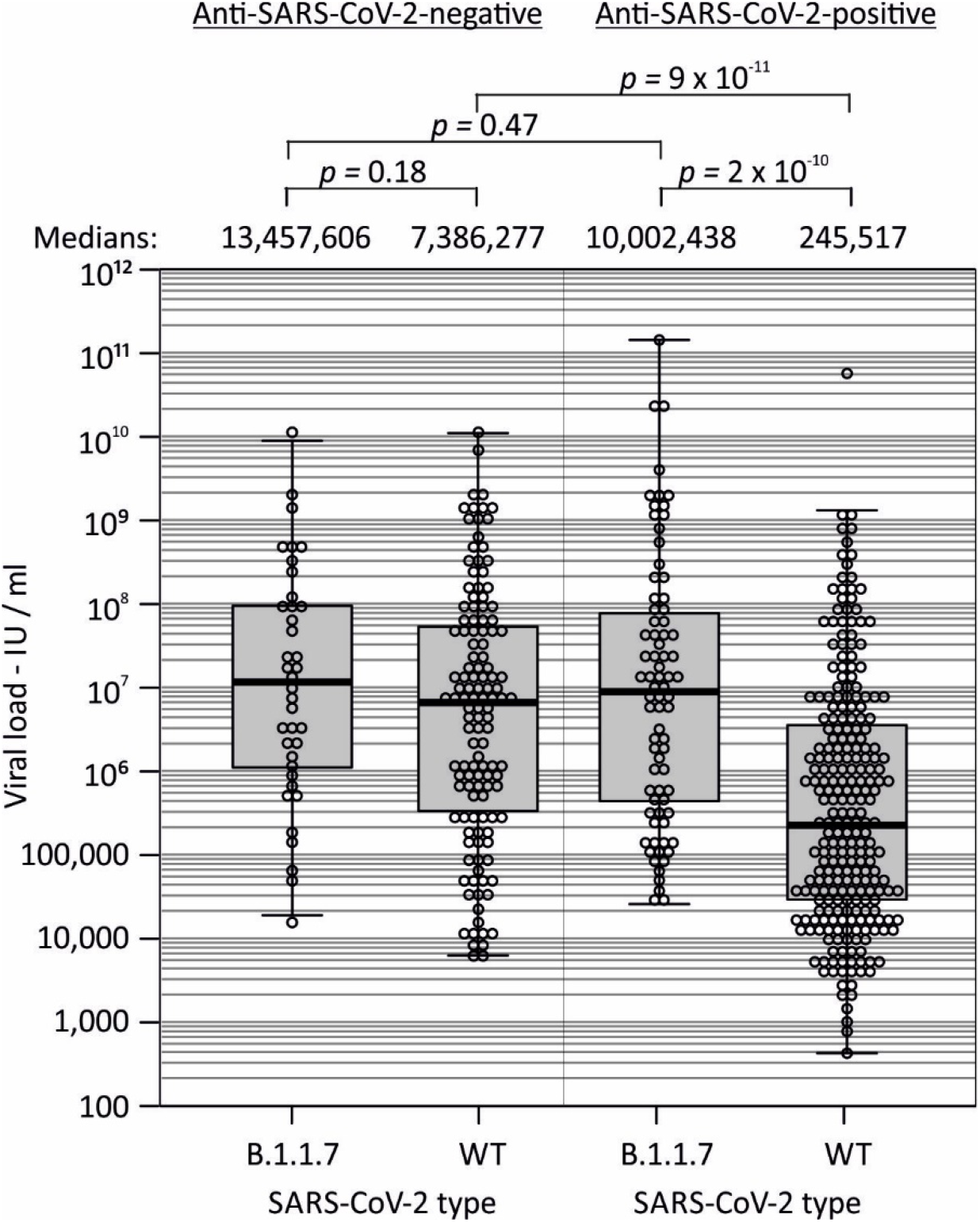
VIRAL LOAD OF WILD-TYPE AND B.1.1.7 STRAINS IN SEROPOSITIVE AND SEROGENATIVE SUBJECTS Distributions of viral loads of in samples from patients infected with wild-type and B.1.1.7 strains, sub-divided by serostatus. Distributions were compared by Kruskall-Wallis non-parametric test.

## DISCUSSION

This is the first study to demonstrate that median viral loads were substantially higher (45-fold) in seronegative COVID-19 patients admitted to the intensive care unit compared to seropositives. This difference was almost entirely due to 40-fold higher median viral loads in seropositive B.1.1.7-infected subjects compared to seropositive wild type -infected subjects. These data are consistent with different kinetics of viral growth, removal, and seroconversion in B.1.1.7-infection compared to wild-type SARS-CoV-2 infection. Our findings suggest that sub-group analysis of therapeutic trials for COVID-19 should examine outcomes not only by viral load and the presence or absence of anti-spike antibody, but also according to viral genotype that may influence the observed treatment efficacy of convalescent plasma.

The motivation for this study was the need to measure virological variables that can influence the efficacy of convalescent plasma therapy, including the emerging new SARS-CoV-2 variants with potentially reduced neutralisation capacity^11,16,19^ and greater pathogenicity^15,20^. Such analyses have been rarely performed in previous or ongoing COVID-19 treatment trials, yet the findings in this severely unwell patient cohort indicate that the studied patients were virologically highly heterogeneous.

Viral loads were determined using quantitative RT-PCR and results calibrated into international units; the use of the NIBSC international standard to express quantitative data using a common standard allows comparison of viral load data obtained in this and independent studies. As previously mentioned, the often reported Ct values provide only an indirect metric of viral load, differing for example through varying efficiencies of primer/probe combinations^21^, which implies that quantitative results are not directly comparable. Most striking was the observation of an approximately billion-fold variation in SARS-CoV-2 viral loads as evaluated by a sensitive and quantitative real time RT-PCR for the conserved N gene. This range is consistent with previous studies that report transient, high levels of SARS-CoV-2 RNA detection in acute infection in nasopharyngeal samples. However, the predictive value of viral loads for clinical outcomes or treatment response is unclear, with several studies reporting significant^22,23^, minimal^24^ or no^25^ associations with increased severity or mortality from COVID-19.

The extreme range of viral loads we observed is consistent with the kinetics of SARS-CoV-2 secretion in the respiratory tract, where rapid decline following the acute infection stage were observed^23,25^. Indeed, the observation that RNA levels were <100 IU/ml in 10% of subjects on PCR screening (Fig. 1A) despite severe COVID-19 leading to ICU admission suggests that a substantial element in the disease mechanisms of this latter group is inflammatory or cytokine-related, rather than being directly virally induced. The higher frequency of seropositivity for SARS-CoV-2 in subjects who were classed as PCR-negative or showed lower viral loads in those infected with the virus with the wild-type genotype (Fig. 1B) provides further evidence for differences in the timings of severe disease presentations relative to the evolution of SARS-CoV-2 infection. Viral loads and anti-SARS-CoV-2 antibody status may therefore be additionally determined by the timing of ICU admission relative to disease onset rather than just intrinsic variability in virus replication between individuals in those infected with the virus with the wild-type genotype.

Prospectively, evaluation of disease stage by viral load measurements and serological status provides potentially valuable information for patient stratification in treatment selection for COVID-19. Antiviral effect of neutralising antibodies in convalescent plasma may be most appropriately employed for patients in early disease stages where respiratory tract pathology is primarily virus-driven. Conversely, those presenting late post-seroconversion may therefore not be helped by infusion of additional neutralising antibodies where disease mechanisms may primarily derive from inflammation-mediated damage and who may better respond to interventions that modulate host response (corticosteroids, IL-6 receptor antagonists^1,26^). The importance of identifying those at early stages of infection is demonstrated by the efficacy of convalescent plasma given within 3 days from disease onset^6^ whereas no benefit was seen when plasma was used in later stage infections (median 8 days from onset^27^).

The effects of mutations in the spike gene on the transmissibility, viral loads, disease severity and potential antigenic variation is currently an area of substantial concern for COVID-19 pandemic management, public health measures and immunisation programmes. SARS-CoV-2 strains in clade B.1.1.7 have become increasingly prevalent in the UK population whilst the REMAP-CAP convalescent plasma trial patients were drawn (Fig. 2). The spike mutations may lead to a reduction in susceptibility to neutralisation by antibodies elicited by infection or immunisation with previously circulating variants of SARS-CoV-2^19,28^.

The development of practical methods for rapid and large-scale identification of B.1.1.7 strains is therefore of paramount importance for monitoring its spread worldwide, given its greater propensity to transmit^14^ and overcome containment measures put in place for previously circulating strains of SARS-CoV-2. The RFLP assay furthermore enables typing to be done with simple laboratory equipment in resource-limited diagnostic facilities without access to HTS or real-time PCR equipment.

The study subjects showed a substantial representation of B.1.1.7 infections, particularly those recruited towards the end of the study period (Fig. 2), mirroring the appearance of this variant in the wider UK population over this period among primarily non-hospitalised individuals. Very recent investigations have provided evidence that B.1.1.7 infections show a 1.7-fold higher hazard of death within 28 day of diagnosis^20^. These findings are consistent with some, but not all, ongoing studies (summarised in^15^), including higher relative case fatality ratios between B.1.1.7 and WT of 1.29-1.36 (Imperial College London) and mortality hazard ratios of 1.7 (PHE) and 1.7 (University of Exeter). In this context, the finding of substantially higher median viral loads in B.1.1.7 infections compared to wild-type in the REMAP-CAP cohort is of substantial mechanistic relevance. The observation that viral loads were higher in B.1.1.7 only in those who had already seroconverted for anti-SARS-CoV-2 antibody (x40-fold; Fig. 4) are consistent with replication capacity and/or a less effective cellular or humoral host immune response to contain B.1.1.7 replication than wild-type. This contrasts with the G614 spike mutant that manifested greater in vitro fitness and replicative capacity compared to wild-type strains with D614^29^. The further variable that may influence the efficacy of convalescent plasma therapy is antigenic variation; the possibility that B.1.1.7-associated amino acid changes in the spike gene may lead to greater resistance to neutralisation is supported by observations of reduced neutralisation susceptibility of B.1.1.7 pseudotypes compared to wild type in some^19^ although not all^30^ studies.

Virus strain characterisation contributes substantially to the virological characterisation of the REMAP-CAP cohort and its association with higher viral loads may potently influence treatment response. While current indications from the REMAP-CAP trial demonstrate no overall effect on the treated cohort compared to untreated controls^10^, data obtained in the current results will form the basis for further analyses of subgroups defined by antibody status, viral load band and by infecting strain. While this analysis is post hoc, findings may identify specific patient groups who will benefit from convalescent plasma and related immunotherapies in the future.

## Supporting information

REMAP-CAP Immunoglobulin Domain UK Investigators

## Data Availability

Anonymised virological data presented in this study is available upon requests directed to the corresponding author. Proposals will be reviewed and approved by the sponsor, investigator, and collaborators on the basis of scientific merit. After approval of a proposal, data can be shared through a secure online platform after signing a data access agreement. Study protocol defining the baseline sampling can be accessed via https://static1.squarespace.com/static/5cde3c7d9a69340001d79ffe/t/5f5c842bd7bf523730bdda76/1599898680445/REMAP-CAP+COVID-19+Immunoglobulin+Therapy+Domain-Specific+Appendix+V1.01-+01+June+2020.pdf.

