## Supplementary Methods and Data for "Virological and serological characterization of critically ill patients with COVID-19 in the UK: a special focus on variant detection"

SUPPLEMENTARY DATA

PROTOCOLS FOR SARS-CoV-2 GENOMIC CHARACTERISATION BY

- Allele-specific probes,
- Restriction fragment length polymorphism assays
- High throughput sequencing

**A) PCR Allele-Specific Probe (PCR-ASP)**

*Differentiation of B.1.1.7 and wt amplicon sequences.* An RT-PCR using allele-specific probes (ASP) for the G→C polymorphism at position 24814 (D1118H) was used to differentiate WT and B.1.1.7 strains. Probe_WT and Probe_B.1.1.7 differ in the SNP at position 24814 but are otherwise identical in sequence (Table S1) – the SNP disrupts probe annealing, resulting in a decrease in the probe $T_M$. When the RT-PCR reaction is performed close to the $T_M$ of the probe, differential binding and amplification of the two probe oligonucleotides occurs dependent on the sequence of the target. The impact of the SNP on binding dynamics is difficult to predict bioinformatically and should be optimized for salt and primer concentrations in the reaction master mix.

*PCR.* For this multiplex reaction, the oligonucleotide working concentrations were 600 nM for Forward and Reverse Primers and 150 nM for Probe_WT and Probe B.1.1.7 (Table S1). Single-use aliquots at 5X the working concentration were prepared upon oligonucleotide resuspension in ultrapure $H_2O$. The Quantitect RT-PCR kit (Qiagen) was used to perform RT-PCR. A 20 µL reaction mix was prepared containing 12.5 µL of RT-PCR Master Mix, 0.25 µL of reverse transcriptase (RT) from the kit, 5 µL of 5X oligo mix, and 2.25 µL of water and was mixed with 5 µL of RNA extract in a MicroAmp Fast Optical 96-well reaction plate (ThermoFisher Scientific). The PCR-ASP assay was carried out on an Applied Biosystems StepOnePlus Real-Time PCR system (ThermoFisher Scientific) on the genotyping program with the following settings: 50°C for 30m (reverse transcription), 60°C for 1m (pre-amplification read), 95°C for 15m, and 45 cycles of 94°C for 15s, 55°C for 20s, and 60°C for 1m, and 60°C for 30s (post-amplification read).

*Results interpretation.* Sequence determination is based on the relative change in probe signal in pre- and post-amplification reads for both probes and can be visualized on a cartesian plane (Figure S1A). From a validation experiment of 40 samples typed according to HTS, an in-house algorithm was derived for PCR-ASP typing (equation 1). Those with ΔProbe_B.1.1.7 > Y were typed as “B.1.1.7”; those with ΔProbe_B.1.1.7 < Y were typed as “WT”. This equation is visualized as a straight line on figure S1A.

**Equation 1**

$$Y = 0.035 * (\Delta Probe_{WT}) + 0.02185$$

Samples that failed to produce ΔProbe_WT of at least 0.06 were determined to be amplification failures and typed ‘inconclusive’. Probe_WT was used for quality control as the FAM-label is brighter than the TAMRA-label.

In this instance, Probe_B.1.1.7 was TAMRA-labelled to leverage the four channels available on the StepOnePlus Real-Time PCR system. However, users with more limited PCR systems may consider selecting VIC or JOE labelling or an equivalent for Probe_B.1.1.7, with the caveat that these combinations have not been evaluated for cross-talk in this assay.

**B) Nested PCR-Restriction Fragment Length Polymorphism (Nested-RFLP)**

A nested PCR-Restriction fragment length polymorphism (Nested-RFLP) was additionally established for B.1.1.7 identification, based on the G→C polymorphism at position 24814 disrupting a *Sfc*I restriction site (5’-CTRYAG-3’). 5 µL of viral RNA was reverse transcribed into cDNA using the SuperScript III Reverse Transcriptase (Thermofisher Scientific) and the 1118 outer primers (Table S1) according to the manufacturer’s protocol for gene-specific primers.

*1st round amplification*, A 25 µL reaction mix containing 2 µL of cDNA, 5 µL of 5X GoTaq Green Master Mix (Promega), 0.125 µL of 5u/µL GoTaq G2 polymerase (Promega), 2 µL of 2.5 mM dNTP mix (Stratech Scientific), 5 µL of 2.5 µM 1118 outer primer mix, and 11.875 µL of PCR-grade water was amplified on thermal cycler with the following settings: 95°C for 2m, 30 cycles of 95°C for 20s, 47°C for 20s, and 72°C for 45s, and a final extension of 72°C for 5m.

*2nd round amplification* A 25 µL reaction mix containing 1 µL of 1st round product, 5 µL of 5X GoTaq Master Mix (Promega), 0.125 µL of 5u/µL GoTaq G2 polymerase (Promega), 2 µL of 2.5 mM dNTP mix (Stratech Scientific), 5 µL of 5X 1118 forward and reverse primer mix, and 12.875 µL of PCR-grade water was amplified on a thermal cycler with the following settings: 95°C for 2m, 30 cycles of 95°C for 20s, 56°C for 20s, and 72°C for 45s, and a final extension of 72°C for 5m. 6 µL of the 2nd round product was digested using *SfcI* (New England Biolabs) according to the manufacturer’s instructions.

*RFLP*. PCR product was visualized by gel electrophoresis in a 2.5% agarose gel containing Sybr-Safe DNA Gel Stain (ThermoFisher Scientific). Uncut amplicons (lineage B.1.1.7) ran near 200 bp and cut amplicons (WT) ran near 120 bp (Figure S1B).

*Specificity.* This assay has the caveat of additional point mutations in the restriction site leading to false positive typing of samples as lineage B.1.1.7. Sample 110 was incorrectly identified as B.1.1.7 by the nested-RFLP; sanger sequencing of the second round product revealed a T→C polymorphism at position 24810 that also ablated the restriction site. This mutation is expected in 2.6% of circulating WT virus in the UK in November 2020 based on GISAID data.

C) **High throughput sequencing[1].** A total of 30 µl of extracted RNA was concentrated to 2 µl using RNAClean XP beads (Beckman) at 1.8 ratio (1.8x) of the RNA starting volume. The SMARTer Stranded Total RNA-Seq Kit v2 (Pico mammalian input; Takara BioSciences) was used to construct total nucleic libraries, following the manufacturer's instructions without the RNA fragmentation or Ribo-depletion steps. The SMARTer Stranded kit uses random primers, tailed with the Illumina read 1 sequence, to start reverse transcription, and a Template Switching Oligo (TSO) to add the Illumina read 2 sequence at the 3' of the synthesized cDNA.

The 1$^{st}$ stranded cDNA was PCR amplified with 12 cycles, using indexed primers, to generate tagged Illumina libraries. Indexed libraries were pooled at equal volume and captured using 120-mer oligonucleotide xGen lockdown probes (Integrated DNA Technologies; IDT) designed in-house to match 60 nt sliding windows of the NC_045512 reference strain. Hybridization of 500 ng of up to 96 pooled DNA libraries was performed overnight with the probes, 5 µg of Cot-1 DNA and 2 µl xGen Universal blocker-TS mix (IDT) using the SeqCap EZ Hybridization and Wash Kit (Roche).

Biotinylated probes were bound to streptavidin M-270 Dynabeads, and virus enriched libraries were amplified with further 12 cycles of PCR using Post-LM Primers 1&2 (Roche). Captured pools were cleaned with 0.68x volume Ampure XP beads (Beckman), and up to 4 pools were combined with equal molarity and sequenced on a single lane of an Illumina SP500 flow cell, using v1.0 chemistry, on an Illumina NovaSeq 6000 system.

**Phylogenetic analysis.** Consensus sequences were aligned in SSE[2]; a phylogenetic tree of those showing greater than 65% coverage was constructed in MEGA7.0[3] using p (uncorrected) nucleotide distances and bootstrap re-sampling. The following reference sequences were included in the tree: prototype SARS-CoV-2 MN908947/2019.95/CN; G: hCoV-19/USA/CA-LACPHL-AF00165/2021, GH: hCoV-19/USA/CA-LACPHL-AF00166/2021, GR: hCoV-19/USA/CA-LACPHL-AF00159/2021, GV: hCoV-19/Japan/IC-0710/2021, L: hCoV-19/Indonesia/BT-LIPI-006/2020, O: hCoV-19/Indonesia/JK-NIHRD-C0023408/2020, S: hCoV-19/France/Bre-IPP11111/2020, V: hCoV-19/USA/NY-AECOM_075/2020,


1. Bonsall D, Golubchik T, de Cesare M, et al. A Comprehensive Genomics Solution for HIV Surveillance and Clinical Monitoring in Low-Income Settings. *J Clin Microbiol* 2020; **58**(10).
2. Simmonds P. SSE: a nucleotide and amino acid sequence analysis platform. *BMCResNotes* 2012; **5**(1): 50.

3. Kumar S, Stecher G, Tamura K. MEGA7: Molecular Evolutionary Genetics Analysis Version 7.0 for Bigger Datasets. *Mol Biol Evol* 2016; **33**(7): 1870-4.

TABLE S1

PRIMERS AND PROBES USED FOR B.1.1.7 TYPING ASSAYS

*Allele-specific PCR:*

<table>
<tr><th>Primer</th><th>Position[1]</th><th>Sequence</th></tr>
<tr><td>Forward</td><td>24745</td><td>5’- GCCATTTGTCATGATGGAAAAG -3’</td></tr>
<tr><td>Reverse</td><td>25002</td><td>5’- CTAATTCAGGTTGCAAAGGATC -3’</td></tr>
<tr><td></td><td></td><td></td></tr>
<tr><td>Probe_WT[2]</td><td>24896</td><td>5’- CCACAAATCATTACTACA|<u>G</u>|ACAACACATTTGTGT-3’</td></tr>
<tr><td>Probe_B.1.1.7[3]</td><td>24896</td><td>5’- CCACAAATCATTACTACA|<u>C</u>|ACAACACATTTGTGT-3’</td></tr>
</table>

*Nested PCR / RFLP:*

| | | |
|---|---|---|
| Outer_forward: | 24632 | 5’- CTTGCTGCTACTAAAATGTCAG-3’ |
| Outer_reverse: | 25184 | 5’- GCTCATACTTTCCAAGTTCTTG-3’ |

[1]Position in the Wuhan-1 prototype sequence (Accession number MN908947)

[2]FAM-labelled; polymorphic site for D1118H at position 24914 shown in bold

[3]TAMRA-labelled; polymorphic site for D1118H at position 24914 shown in bold

TABLE S2

FREQUENCIES OF SPIKE DELETION H69 & V70 OVER STUDY PERIOD

| **Location / Period** | **Strain** | **Total** | **Insert** | **Del** | **Specificity** |
|---|---|---|---|---|---|
| All | B.1.1.28 | 374 | 374 | 0 | 100.00% |
| All | B.1.351 | 834 | 834 | 0 | 100.00% |
| UK_Dec-2020 | B.1.1.7 | 5636 | 0 | 5636 | 100.00% |
| UK_Jan-2021 | B.1.1.7 | 20686 | 0 | 20686 | 100.00% |
| UK_Nov-2020 | B.1.1.7 | 6 | 0 | 6 | 100.00% |
| UK_Jan-2021 | WT | 4014 | 3982 | 32 | 99.20% |
| UK_Dec-2020 | WT | 9852 | 9654 | 198 | 97.99% |
| UK_Nov-2020 | WT | 9259 | 9020 | 239 | 97.42% |
| UK_July-2020 | WT | 6961 | 6960 | 1 | 99.99% |
| UK_Jan-April-2020 | WT | 6961 | 6961 | 0 | 100.00% |

TABLE S3

CONCORDANCE OF ASP AND RFLP ASSAYS WITH HTS

| | | | ASP | | | RFLP | | |
|---|---|---|---|---|---|---|---|---|
| | **HTS** | **Total** | **WT** | **B.1.1.7** | **NR**[1] | **WT** | **B.1.1.7** | **NR**[1] |
| | WT | 30 | 30 | 9 | 0 | 26 | 1[2] | 3 |
| | B.1.1.7 | 10 | 0 | 9 | 1 | 0 | 9 | 1 |

[1]No result obtained

[2]Sanger sequencing identified a T→C polymorphism at position 24810 that disrupted the restriction site

FIGURE S1

A)

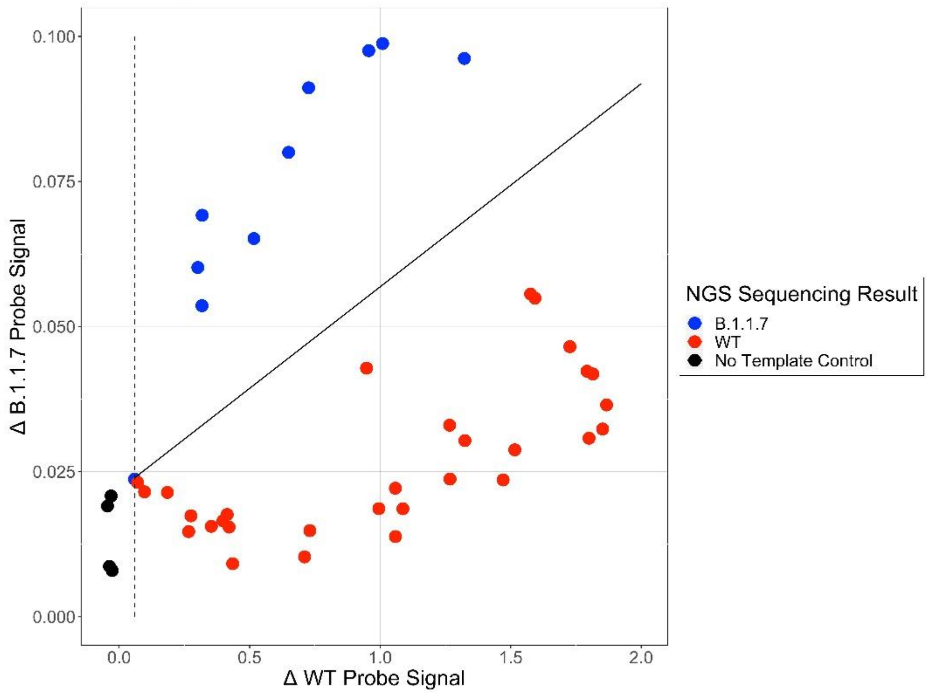


B)

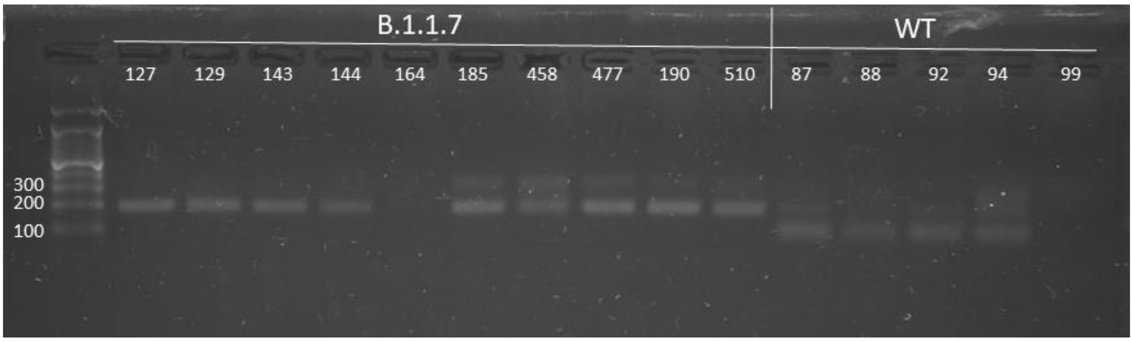


A) Hybridization scores of amplified DNA sequences from WT and B.1.1.7 SARS-CoV-2 strains to FAM-WT and TAMRA-B.1.1.7 labelled probes (Table S1; Suppl. Data) for type identification. (B) Representative agarose gel of PCR amplicons cleaved with *Sfc*I. The G->C change underlying the D1118H mutation disrupted the *Sfc*I restriction site and is uncleaved (199 bp product – left 10 lanes) while the WT is cleaved (~120 bp product – right 5 lanes). Samples 164 and 99 did not produce nested PCR products. Bands running ~550 bp are contaminating first round product; WT sequences may show incomplete digestion.

FIGURE S2

RELATIONSHIP OF SEQUENCING COVERAGE TO SAMPLE VIRAL LOAD

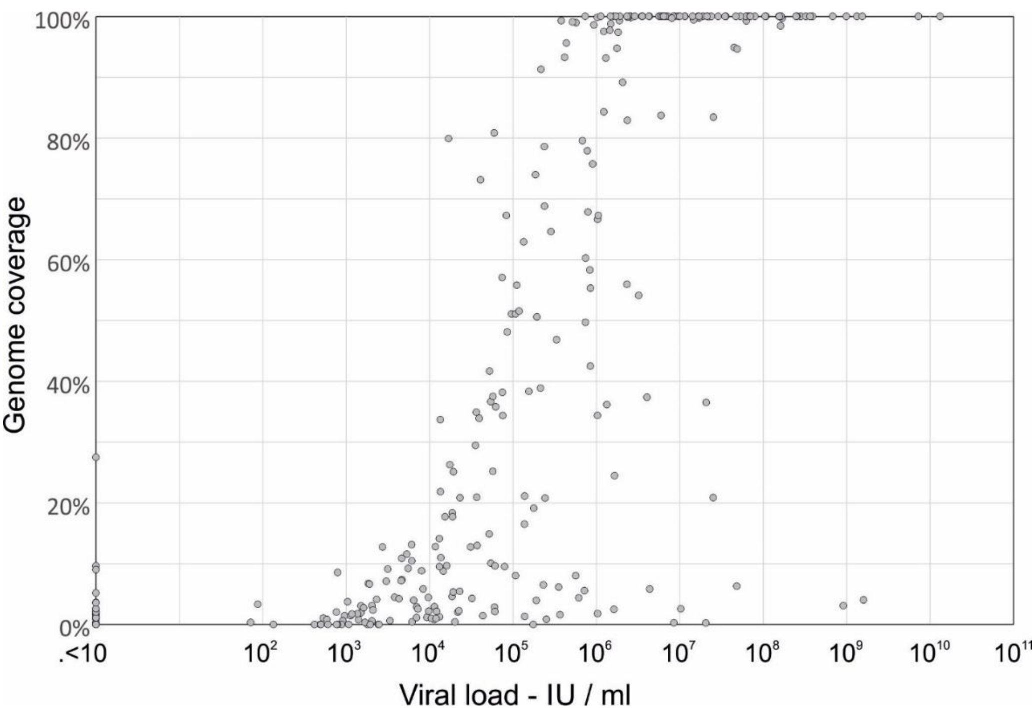


Genome coverage of consensus sequences of SARS-CoV-2 in study samples.

FIGURE S3

PHYLOGENETIC ANALYSIS OF SARS-CoV-2 SEQUENCES FROM STUDY SUBJECTS

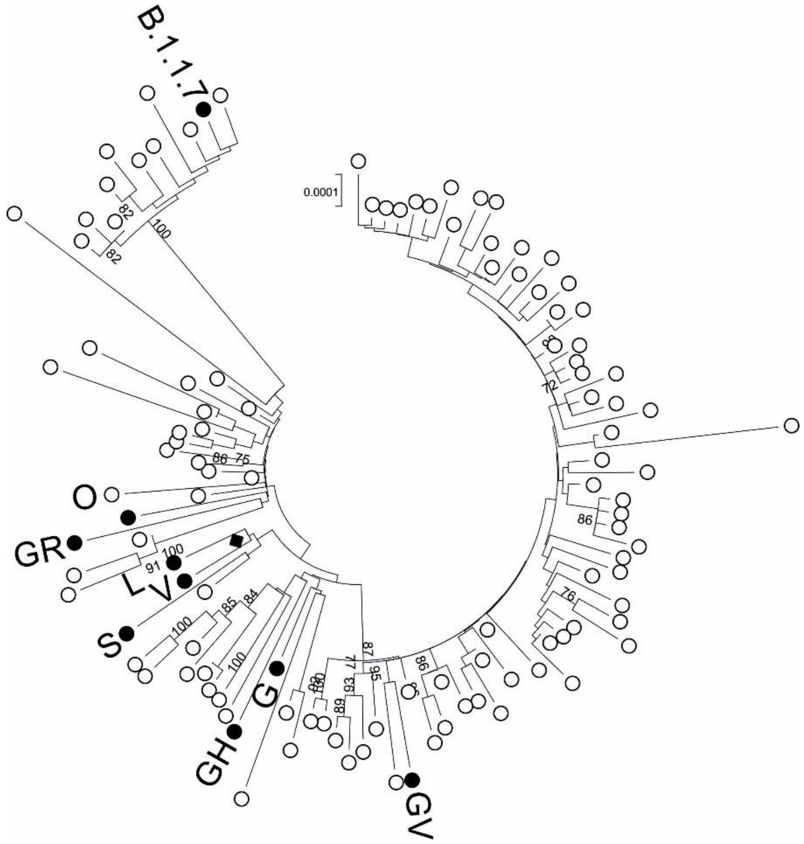


Neighbour-joining tree of SARS-CoV-2 sequences obtained from the study subjects (restricted to those >65% complete). For clade identification, representative reference sequences of each of the clades defined by GISAID were included. The black diamond in the sequences of thew Wuhan-1 prototype strain. The majority clade (GV) corresponds to B.1.1.7 (Rambaut *et al.,* 2021) or 20E (NextStrain). Bootstrap support (100 replicates) indicated on branches (values >70% shown).

FIGURE S4

SUCCESS RATE OF TYPING ASSAYS FOR SAMPLES OF DIFFERENT VIRAL LOADS

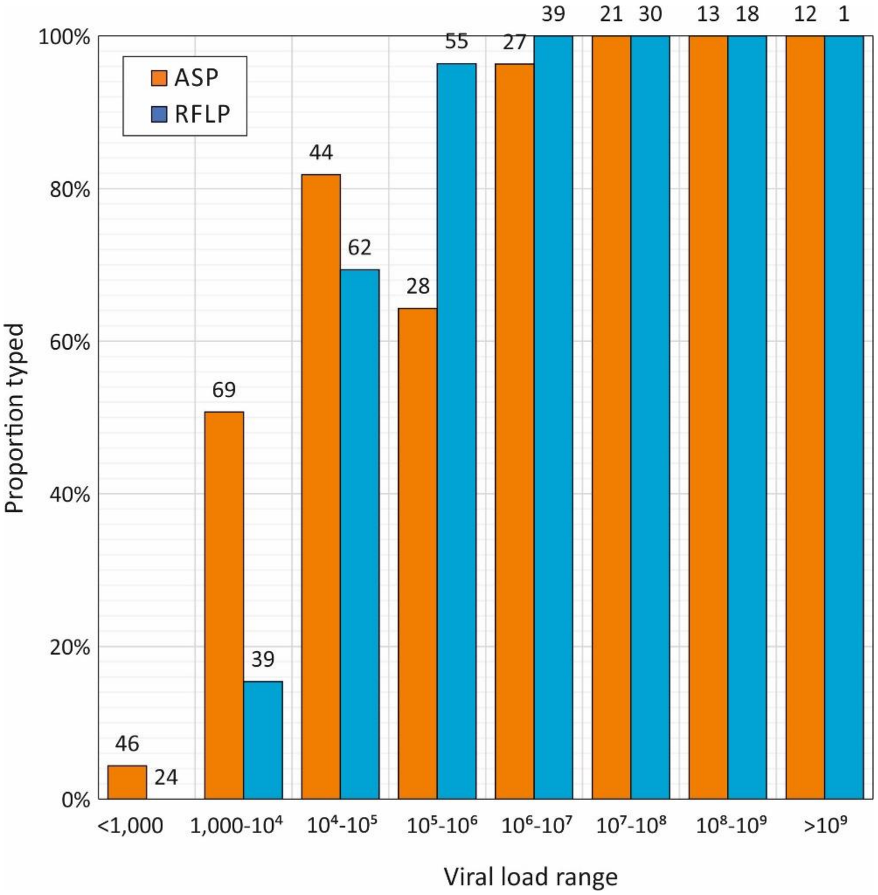
