## Supplementary material for "Virological and serological characterization of critically ill patients with COVID-19 in the UK: a special focus on variant detection": REMAP-CAP Immunoglobulin Domain UK Investigators

### *REMAP-CAP Trial Investigators & Collaborators associated with this sub-study*

**International Trial Steering Committee:**
Farah Al-Beidh, Derek Angus, Djillali Annane, Yaseen Arabi, Abi Beane, Wilma van Bentum-Puijk, Scott Berry, Zahra Bhimani, Marc Bonten, Charlotte Bradbury, Frank Brunkhorst, Meredith Buxton, Allen Cheng, Lennie Derde, Lise Estcourt, Herman Goossens, Anthony Gordon, Cameron Green, Rashan Haniffa, Francois Lamontagne, Patrick Lawler, Edward Litton, John Marshall, Colin McArthur, Daniel McAuley, Shay McGuinness, Bryan McVerry, Stephanie Montgomery, Paul Mouncey, Srinivas Murthy, Alistair Nichol, Rachael Parke, Jane Parker, Kathryn Rowan, Marlene Santos, Christopher Seymour, Alexis Turgeon, Anne Turner, Frank van de Veerdonk, Steve Webb (Chair), Ryan Zarychanski

**Regional Management Committees:**
***European Regional Management Committee***
Farah Al-Beidh, Derek Angus, Djillali Annane, Wilma van Bentum-Puijk, Scott Berry, Marc Bonten (Executive Director and Chair), Nicole Brillinger, Frank Brunkhorst, Maurizio Cecconi, Lennie Derde, Stephan Ermann, Bruno Francois, Herman Goossens, Anthony Gordon, Cameron Green, Sebastiaan Hullegie, Rene Markgraff, Colin McArthur, Paul Mouncey, Alistair Nichol, Mathias Pletz, Pedro Povoa, Gernot Rohde, Kathryn Rowan, Steve Webb

**Regional Coordinating Centers:**
***Europe:*** University Medical Center Utrecht (UMCU)
***United Kingdom:*** Intensive Care National Audit and Research Centre (ICNARC), and Imperial College London

**COVID-19 Immunoglobulin Domain-Specific Working Group:**
Derek Angus, Donald Arnold, Phillipe Begin, Scott Berry, Richard Charlewood, Michael Chasse, Mark Coyne, Jamie Cooper, James Daly, Lise Estcourt (Chair, UK lead), Dean Fergusson, Anthony Gordon, Iain Gosbell, Heli Harvala, Tom Hills (New Zealand lead), Christopher Horvat, David Huang, Sheila MacLennan, John Marshall, Colin McArthur (New Zealand lead), Bryan McVerry (USA lead), David Menon, Susan Morpeth, Paul Mouncey, Srinivas Murthy, John McDyer, Zoe McQuilten (Australia lead), Alistair Nichol (Ireland lead), Nicole Pridee, David Roberts, Kathryn Rowan, Christopher Seymour, Manu Shankar-Hari (UK lead), Helen Thomas, Alan Tinmouth, Darrell Triulzi, Alexis Turgeon (Canada lead), Tim Walsh, Steve Webb, Erica Wood, Ryan Zarychanski (Canada lead)

**Statistical Analysis Committee:**
Michelle Detry, PhD, Mark Fitzgerald, PhD, Roger Lewis, MD, PhD (Chair), Anna McGlothlin, PhD, Ashish Sanil, PhD, Christina Saunders, PhD

**Statistical Design Team:**
Lindsay Berry, PhD, Scott Berry, PhD, Elizabeth Lorenzi, PhD

**Project Management:**
***Europe:*** Wilma van Bentum Puijk, Wietske Bouwman, Radhika Ganpat, Erika Groenveld, Denise van Hout, Yara Mangindaan, Clementina Okundaye, Lorraine Parker, Svenja Peters, Ilse Rietveld, Linda Rikkert, Kik Raymakers, Irma Scheepstra-Beukers, Albertine Smit,
***Global:*** Cameron Green

***United Kingdom:*** Farah Al-Beidh, Aisha Anjum, Janis-Best Lane, Elizbeth Fagbodun, Lorna Miller, Paul Mouncey, Karen Parry-Billings, Sam Peters, Alvin Richards-Belle, Michelle Saull, Stefan Sprinkmoller, Daisy Wiley

**Data and Safety Monitoring Board:**
Julian Bion, Jason Connor, (Deputy Chair), Simon Gates, Victoria Manax (Chair), Tom van der Poll, John Reynolds

**Clinical Trials Groups:**
The REMAP-CAP platform is supported by the Australian and New Zealand Intensive Care Society Clinical Trials Group, the Canadian Critical Care Clinical Trials Group, the Irish Critical Care Clinical Trials Network, the UK Critical Care Research Group and the International Forum of Acute Care Trialists.

REMAP-CAP was supported in the UK by the NIHR Clinical Research Network and we acknowledge the contribution of Kate Gilmour, BSc (Hons), Karen Pearson, MSc, Chris Siewerski, MSc, Sally-Anne Hurford, MSc, Emma Marsh, FdSC, Debbie Campbell, Penny Williams, MSc, Kim Shirley, LLB Hons, Meg Logan, NVQ, Jane Hanson, Becky Dilley, BSc, Louise Phillips, CIM, Anne Oliver, MSc, Mihaela Sutu, MSc, Sheenagh Murphy, PGDip, Latha Aravindan, PhD, Joanne Collins, MRes, Holly Monaghan, Adam Unsworth, NVQ, Seonaid Beddows, MSc, Laura Ann Dawson, LLM, Sarah Dyas, Adeeba Asghar, MSc, Kate Donaldson, BA, Tabitha Skinner, BSc, Nhlanhla Mguni, BSc (Hons), Natasha Muzengi, BSc, Ji Luo, PhD, Joanna O'Reilly, BSc (Hons), Chris Levett, MSc, Alison Potter, David Porter, PhD, Teresa Lockett, MSc, Jazz Bartholomew, MSc, Clare Rook, MSc, Rebecca McKay, Hannah Williams, MSc, Alistair Hall, FRCP, Hilary Campbell, BSc (Hons), Holly Speight, BA, Sandra Halden, Susan Harrison, Mobeena Naz, Kaatje Lomme, MA, Paula Sharratt, MSc, Johnathan Sheffield, FRCP, William Van't Hoff, FRCPCH, James D Williamson, PhD, Alex Barnard, BSc (Hons), Catherine Birch, BA, Morwenna Brend, PhD, Emma Chambers, PhD, Sarah Crawshaw, PhD, Chelsea Drake, BSc, Hayley Duckles-Leech, PhD, Justin Graham, BSc (Hons), Heather Harper, BSc (Hons), Stephen Lock, PGCert, Nicola McMillan, PhD, Cliodhna O'Flaherty, Eleanor OKell, PhD, Amber Hayes, Sally Sam, BSc, Heather Slade, MSc, Susan Walker, PhD, Karen Wilding, MRes, Jayne Goodwin, Helen Hodgson, Yvette Ellis, Dawn Williamson, Madeleine Bayne, MSc, Shane Jackson, Rahim Byrne, Sonia McKenna, Alsion Clinton, NIHR Urgent Public Health Group: https://www.nihr.ac.uk/documents/urgent-public-health-group-members/24638#Members

**Site Investigators and Research Coordinators:**

**Convalescent Plasma Clinical Trials Unit (on behalf of the UK Blood Services):**
L Estcourt (Lead), E Arbon, A Deary, F Clemons, N Dallas, A Evans, C Fitzpatrick-Creamer, C Foley, R Hodge, C Hudson, K Keen, E Laing, M Lobo-Clarke, J Mullings, R Paul, G Powter, S Shanmugaranjan, S Sweity and all staff at NHSBT Clinical Trials Unit that have worked on the Convalescent Plasma Project.

**Oxford Bioarchive team:** David Roberts, Rutger Ploeg, Hoi Pat Tsang, Marta Oliveira, Sheba Ziyenge, Ullrich Leuschner, Nick Ciccone, Sarah Cross and all staff at Oxford Bioarchive team

**Shankar-Hari Group:** Matthew Fish, Jennifer Rhynne, Aislinn Jennings, Jakob Jeriha, Silvia Cellone Trevelin, Isabella Tosi, James Dylan Williams, Carolyn Karman Lam and Rashida Pramanik (https://www.kcl.ac.uk/research/shankar-hari-group)

***United Kingdom***:

*Aberdeen Royal Infirmary:* Callum Kaye, MBChB, Angela Allan, PGDip; Transfusion: J Lussier, M Mathie, L Jappy and all in the Blood Bank Laboratory.

*Addenbrooke's Hospital:* Charlotte Summers, PhD, Petra Polgarova MSc; Transfusion: K Philpott, C Newsam, M Lewin, T Moore, H Madiyiko, V Rose, S Grist, R Smith, H Dakers-Black, M Ibrahim and the laboratory team.

*Alder Hey Children's NHS Foundation Trust:* Stephen J McWilliam, PhD, Daniel B Hawcutt, MD, Laura Rad, BSc(Hons), Laura O'Malley, BSc(Hons), Jennifer Whitbread, BSc(Hons); Transfusion: T Shackleton, S Owens, J Fu.

*Alexandra Hospital Redditch:* Olivia Kelsall, MBChB, Nicholas Cowley MD, Laura Wild, BSc(Hons), Jessica Thrush, RGN, Hannah Wood, BSc(Hons), Karen Austin, RGN; Transfusion: C Khan, G Godding, E Murphy, E Loxley and the Blood Transfusion laboratory team.

*Altnagelvin Hospital:* Adrian Donnelly, FFICM, Martin Kelly, MD, Naoise Smyth MB ChB, Sinéad O'Kane, BSc(Hons), Declan McClintock, MSc, Majella Warnock, MPharm, Ryan Campbell BSc, Edmund McCallion MPharm; Transfusion: A Crawford, B O'Neill, M P McNicholl, J Monaghan, N Smyth, C Kelly, C Kerlin and Haematology/Blood Transfusion/Central Specimen Reception laboratory teams.

*Antrim Area Hospital:* Paul Johnson, FFARCSI, Shirley McKenna, MSc, Joanne Hanley, MSc, Andrew Currie, MSc, Barbara Allen, MPharm, Clare Mc Goldrick, MPhil, Moyra Mc Master, RGN, ; Transfusion: C A Henry, B Graham, K Potter and the Blood Transfusion laboratory team.

*Barnet Hospital:* Rajeev Jha, MD, Michael Kalogirou, MD, Christine Ellis, PhD, Vinodh Krishnamurthy, PhD, Vashish Deelchand, MSc, Aibhilin O'Connor, MSc; Transfusion: R Atugonza, A Li, J Li, S Tekle, A M Ellis, A Natarajan, S Mellin and the Blood Transfusion laboratory team.

*Basildon Universty Hospital: Dipak Mukherjee, MD, Agilan Kaliappan, MD, Anirudda Pai, MD,Mark Vertue, Anne Nicholson, Joanne Riches, Gracie Maloney, Lauren Kittridge, Amanda Solesbury, Kezia Allen;* Transfusion: T Green, M O'Connell and the Blood Transfusion laboratory team.

*Belfast Health and Social Care Trust (Belfast City Hospital, Mater Infirmorium, Royal Victoria Hospital):* Jon Silversides, PhD, Peter McGuigan, MBBCh, Kathryn Ward, BSc, Aisling O'Neill, BSc, Stephanie Finn, BSc; Transfusion: C A Henry, P Windrum, B O'Neill, S Reilly and Blood Transfusion laboratory team.

*Brighton and Sussex University Hospitals Trust:* Barbara Phillips, Laura Oritz-Ruiz de Gordoa, BSc; Transfusion: J Cole, L Humber and the Blood Transfusion laboratory team.

*Bristol Royal Infirmary:* Jeremy Bewley, MBChB, Matthew Thomas, MBChB, Katie Sweet, BSc(Hons), Lisa Grimmer, BSc(Hons), Rebekah Johnson, BSc(Hons); Transfusion: S White, A Wardle, S Cooke, R Cowburn, A Parejasanchez, L Johnston, S Goolam-Hossen and the Blood Transfusion laboratory teams.

*Calderdale and Huddersfield Foundation Trust:* Jez Pinnell, MD, Matt Robinson, BSc(Hons), Lisa Gledhill, MSc, Tracy Wood, BSc(Hons); Transfusion: S Kershaw, P Desai, M Lake, H Senior and the Blood Transfusion laboratory team.

*Cardiff and Vale University Health Board:* Matt Morgan, PhD, Jade Cole, BSc, Helen Hill, BSc, Michelle Davies, BN, Angharad Williams, BSc, Emma Thomas, BSc, Rhys Davies, BSc, Matt Wise, DPhil; Transfusion: R Carnegie, S McWilliam, A Patterson, R Borrell, C M Neville, L Parkinson, G Andrikopoulos, S Burns and the Blood Transfusion laboratory team.

*Charing Cross Hospital:* David Antcliffe, PhD, Maie Templeton, MSc, Roceld Rojo, BSN, Phoebe Coghlan, MA, Joanna Smee, BSc; Transfusion: A Rahman, D Johnson, L Chapple, E Nweje, K Sabljak, S Kassa, U Munu, H Dawson, F Regan, F Chowdhury, V Chiroma and Blood Transfusion laboratory teams.

*Chesterfield Royal Hospital:* Euan Mackay, MD, Jon Cort, MD, Amanda Whileman, BSc, Thomas Spencer, Nick Spittle, Sarah Beavis, MD, Anand Padmakumar, MD, Katie Dale, BSc, Joanne Hawes, BSc, Emma Moakes, BSc, Rachel Gascoyne, BSc, Kelly Pritchard, BSc, Lesley Stevenson, BSc, Justin Cooke, MD, Karolina Nemeth-Roszpopa, MD; Transfusion: CJ Lattimore, J Smith and the whole transfusion lab team.

*The Christie NHS Foundation Trust:* Vidya Kasipandian, FFICM, Amit Patel, Suzanne Allibone, Roman Mary-Genetu, BSc; Transfusion: D Seals, S Jackson

*Colchester Hospital:* Mohamed Ramali, FRCA, Ooi HC, MRCEM, Alison Ghosh, RN, Rawlings Osagie, PharmD, Malka Jayasinghe Arachchige, MBBS, Melissa Hartley, MBBS; Transfusion: E Byworth, M Mohan, S Turner, A Pereira and the Blood Transfusion laboratory team.

*Countess of Chester Hospital:* Peter Bamford, FFICM, Emily London, MBChB, Kathryn Cawley, MRes, Maria Faulkner, BSc, Helen Jeffrey, DipNS; Transfusion: N Swarbrick, L Hodgkinson and the Blood Transfusion laboratory team

*Croydon University Hospital:* Ashok Sundar Raj, MD, Georgios Tsinaslanidis, MD, Reena Nair Khade, BSc, Gloria Nwajei Agha, BSc, Rose Nalumansi Sekiwala; Transfusion: M Free, M Mbwembwe, S Conran, B Cheung and the Blood Transfusion laboratory team.

*Cumberland Infirmary:* Tim Smith, FRCA, Chris Brewer, BPharm(Hons), Jane Gregory, BSc(Hons); Transfusion: J Sutton, J Nicholson and the Blood Transfusion laboratory team.

*Darlington Memorial Hospital:* James Limb, FRCA, Amanda Cowton, BSc(Hons), Julie O'Brien, DipNurs, Kelly Postlethwaite, DipNurs; Transfusion: J Lawson, C Morrell, S Watson, A Iqbal, R Pipes, J Gilbert and the Blood Transfusion laboratory team.

*Derriford Hospital:* Nikitas Nikitas, PhD, Colin Wells, MSc, Liana Lankester, PGCert, Helen McMillan, MSc; Transfusion: M Binney, C Lowe and the Blood Transfusion laboratory team.

*Dorset County Hospital:* Mark Pulletz, FFICM, Patricia Williams, AdDip, Jenny Birch, BA, Sophie Wiseman, Mpharm, Sarah Horton, BA(Hons); Transfusion: M Greenslade, L Poole, N Dewland, M Deighton and the Blood Transfusion laboratory team.

*East Kent Hospitals (Queen Elizabeth the Queen Mother Hospital):* Ana Alegria, CCT, Salah Turki, MBBch, Tarek Elsefi, MRCP, Nikki Crisp, BSc, Louise Allen, BSc; C Lorenzen, R Spicer, B Rayner, L March, K Kolsteren, L Boorman, R Jennings, H Moore, S Lymn, F Turner and the Blood Transfusion laboratory team.

*East Lancashire Hospitals NHS Trust (Royal Blackburn Hospital):* Matthew Smith, MD, Sri Chukkambotla, MD, Wendy Goddard, BSc, Stephen Duberley BSc; Transfusion: L Mannion, M Sokolowski, S Rigby, T Johnson and Blood Transfusion laboratory team.

*Freeman Hospital and Royal Victoria Infirmary, Newcastle upon Tyne*: Iain J McCullagh, FRCA, Philip Robinson, MSc, Bijal Patel, MSc, Sinead Kelly, PGDip; Transfusion: A Muir, A Baird, S Rowan-Ferry, G Smithson, M Wilson, C Kennedy, M Evans, H Ranton and the Blood Transfusion laboratory team

*Frimley Health NHS Foundation Trust:* Omar Touma, MD, Susan Holland, Christopher Hodge, Holly Taylor, Meera Alderman, Nicky Barnes, Joana Da Rocha, BSc, Catherine Smith, BSc, Nicole Brooks, Thanuja Weerasinghe, BSc, Julie-Ann Sinclair, Yousuf Abusamra, MD, Ronan Doherty, MD, Joanna Cudlipp, MD, Rajeev Singh, MD, Haili Yu, MD, Admad Daebis, MD, Christopher NG, MD, Sara Kendrick, MD, Anita Saran, MD, Ahmed Makky, MD, Danni Greener, MD, Louise Rowe-Leete, Dip, Alexandra Edwards, Dip, Yvonne Bland, BSc, Rozzie Dolman, BSc, Tracy Foster, BSc; Transfusion: N Johnson, J Newanji, J Finden, C Cole, K East, A Kandaswamy and the Blood Transfusion laboratory teams.

*Gateshead Health NHS Trust:* Vanessa Linnett, MD, Amanda Sanderson, Jenny Ritzema, Helen Wild; Transfusion: L Sudlow

*George Eliot Hospital:* Divya Khare, FRCA, Meredith Pinder, BSN, Selvin Selvamoni, MSc, Amitha Gopinath, MBA; Transfusion: T Taylor, E Sharrod, G Roper, R Wookey, L Dyble, K Jawaid, S Chucha, G Ripley, Chaven, R Khan, K Steinert, M Scally, W Penlington and the Blood Transfusion laboratory team.

*Glan Clwyd Hospital:* Richard Pugh, FFICM, Daniel Menzies, FRCP, Richard Lean, MBChB, Xinyi Qiu, MBChB, Jeremy James Scanlon, MBChB; Transfusion: L Hughes, E Hall and the Blood Transfusion laboratory team.

*Glasgow Royal Infirmary:* Kathryn Puxty, MD, Susanne Cathcart, BSc, Chris Mc Govern, MBChB, Samantha Carmichael, MRPharms, Dominic Rimmer, BSc; Transfusion: M Caldwell, A David and the Transfusion Laboratory team.

*Glenfield Hospital Leicester:* Hakeem Yusuff, FFICM, Graziella Isgro, FFICM, Chris Brightling, PhD, Michelle Bourne, BSc(Hons), Michelle Craner, DipHE, Rebecca Boyles, BSc (Hons); Transfusion: A Ghattaoraya, H Qureshi, Yasin Fozdar, T Dinh, M Browett, all UHL Blood Transfusion Laboratory Staff.

*Great Western Hospitals NHS Foundation Trust:* Malcolm Watters, MBBCh, Rachel Prout, MBChB, Louisa Davies, BSc, Suzannah Pegler, BSc(Hons), Lynsey Kyeremeh, BPharm, Aiman Mian, MBBS; Transfusion: J Uppal, S Charlton and the Blood Transfusion team.

*Guy's & St Thomas' NHS Foundation Trust:* Manu Shankar-Hari, PhD, Marlies Ostermann, PhD, Marina Marotti, BSc, Neus Grau Novellas, BSc, Aneta Bociek, BSc; Transfusion: T Maggs, L Woodford, J Wood, J Jones, B Crane, S Turawa, C Lobato, C Furtado, R O'Dea, U Wood, D Jeyapalan and the Blood Transfusion laboratory teams.

*Hammersmith Hospital:* Stephen Brett, MD, Sonia Sousa Arias, BSc, Rebecca Elin Hall, BN; Transfusion: A Rahman, D Johnson, L Chapple, E Nweje, K Sabljak, S Kassa, U Munu, H Dawson, F Regan, F Chowdhury, V Chiroma and Blood Transfusion laboratory teams.

*Homerton University Hospital NHS Foundation Trust:* Susan Jain, MD, Abhinav Gupta MD, Catherine Holbrook; Transfusion: E Fonyonga, A Martinez, M Elmi and the Blood Transfusion laboratory team.

*James Cook University Hospital:* Jeremy Henning, MB, Stephen Bonner, BSc, Keith Hugill, BSc, Emanuel Cirstea, MSc, Dean Wilkinson, BSc, Jessica Jones, BSc; Transfusion: C Elliott, Carolyn Carveth-Marshall and the Blood Transfusion laboratory team.

*James Paget University Hospitals:* Michal Karlikowski, MD, Helen Sutherland, BSc(Hons), Elva Wilhelmsen, DipHE, Jane Woods, BSc, Julie North, BSc(Hons); Transfusion: S Parsons, J Jackson and the Blood Transfusion laboratory team.

*Kettering General Hospital:* Dhinesh Sundaran, FFICM, Laszlo Hollos, FFICM, Susan Coburn, PGCert, Anna Williams, BSc, Samantha Saunders, BTEC; Transfusion: J Frankcam, M Silverstone, A Houston, E Rich, C Little and the Blood Transfusion laboratory team.

*King's College Hospital (Denmark Hill site):* Phil Hopkins, MD, John Smith, RN, Harriet Noble, RN, Maria Theresa Depante, RN, Emma Clarey, RN; Transfusion: K Amenyah, K Nwankiti, J Davies, D Veniard, S Cole, P Gold, C Hawkins and the Blood Transfusion laboratory team.

*Lancashire Teaching Hospitals NHS Foundation Trust:* Shondipon Laha, FFICM, Mark Verlander, MBA, Alexandra Williams, MSc; Transfusion: A Noyon, D Wallbank, S Baines, E Fisher, K Treuberg, B Houghton, E Ly and the Blood Transfusion laboratory team.

*Leeds Teaching Hospitals Trust:* Elankumaran Paramasivam, FRCP, Elizabeth Wilby, BSc (Hons), Bethan Ogg, BSc (Hons), Clare Howcroft, BSc (Hons), Angelique Aspinwall, BSc (Hons), Sam Charlton, BSc (Hons), Richard Gould, MBBS, Deena Mistry, MPharm, Sidra Awan, MPharm, Caroline Bedford, MPharm; Transfusion: A Ballinger, D Howarth, A Liversidge, S Ferguson, J Rock, M Karakantza and the Blood Transfusion laboratory team.

*Leicester General Hospital:* Andrew Hall, MRCP, Jill Cooke, RGN, Caroline Gardiner-Hill, RGN, Carolyn Maloney, Nigel Brunskill, PhD; Transfusion: A Ghattaoraya, H Qureshi, Yasin Fozdar, T Dinh, M Browett, all UHL Blood Transfusion Laboratory Staff.

*Leicester Royal Infirmary:* Hafiz R Qureshi, MRCPI, Neil Flint, MBChB, Sarah Nicholson, Sara Southin, Andrew Nicholson, Amardeep Ghattaoraya, MSc; Transfusion: A Ghattaoraya, H Qureshi, Yasin Fozdar, T Dinh, M Browett, all UHL Blood Transfusion Laboratory Staff.

*Lewisham and Greenwich NHS Trust:* Dr Daniel Harding, MD, Sinead O'Halloran , Amy Collins Emma Smith , Estefania Trues; Transfusion: P Richards, A-M Johnson, T Kelani and laboratory team.

*Liverpool Foundation Trust Aintree:* Barbara Borgatta, PhD, Ian Turner-Bone, DipHE, Amie Reddy, Laura Wilding, DipHE; Transfusion: K Knowles, H Wissett, R Wright, C Dragomir, A Peacock, J Bell, J Gorry, I Houghton, P Burgess and all members of the lab BMS and MLA teams.

*Liverpool Heart and Chest Hospital:* Craig Wilson, Zuhra Surti; Transfusion: K Knowles, H Wissett, R Wright, C Dragomir, A Peacock, J Bell, J Gorry, I Houghton, P Burgess and all members of the lab BMS and MLA teams.

*Luton and Dunstable University Hospital:* Loku Chamara Warnapura, FFICM, Ronan Agno, BSc, Prasannakumari Sathianathan, MSc, Deborah Shaw, FFICM, Nazia Ijaz, FFICM, Dean Burns, MD, Mohammed Nisar, MD, Vanessa Quick, MD, Craig Alexander, BSc, Sanil Patel BSc, Nafisa Hussain, Yvynne Croucher, BSc, Eva-Maria Lang, MD, Banu Rudran, MD, Syed Gilani, MD, Talia Wieder, MD, Margaret Louise Tate, BSc; Transfusion: D Fisher, E Strakosch, Charlotte Alford, M Parmar, Noha Gasmalseed, L Hunold, A Gouveia, S Khan, R Hussein, T Mukwa, G Mmadubuko, J Nnadi, J Dijo, T Jacques- Brown, J Tan and Blood Bank team.

*Maidstone and Tunbridge Wells NHS Trust:* David Golden, FFICM, Miriam Davey, PGDip, Rebecca Seaman BSc (Hons); Transfusion: R Reilly, E Small, W Bonnert, F Ajeneye, C Moore, C Lawler, C Boyd and the Blood Transfusion laboratory team.

*Manchester Royal Infirmary:* Tim Felton, FFICM, Jonathan Bannard-Smith, FFICM, Joanne Henry, Richard Clark, DiPHE, Kathrine Birchall, BSc(Hons), Joanne Henry, MA, Fiona Pomeroy, BSc (Hons), Rachael Quayle, Dip.HE, Katharine Wylie, MSc, Anila Sukuraman, BSc, John McNamarra, MD; Transfusion: T Trimble, E Cooperwaite, C Parker, S Pendlebury, J Peters, S Khan, E Anyanwu, M Evans and the Blood Transfusion laboratory teams.

*Medway Maritime Hospital:* Arystarch Makowski, PhD, Beata Misztal, PhD, Iram Ahmed, PhD, Kevin Neicker, MBA, Sam Millington, BMBS, Rebecca Squires, BSc, Masroor Phulpoto, MBBS; Transfusion: R Nicholas, S Haskins, J Walker, A Davis, S Arnott and the Blood Transfusion laboratory team.

*Milton Keynes University Hospital:* Richard Stewart, Esther Mwaura, BSc, Louise E Mew, BSc(Hons), Lynn Wren, BSc(Hons), Felicity Willams, PhD; Transfusion: J Beharry, T Perry, C Lowe, M Khan, N Jacob and the Blood Transfusion laboratory team.

*Mid & South Essex NHS Foundation Trust:* Aneta Oborska, FRCA, Rino Maeda, MBBS, Selver Kalchko-Veyssal, MD, Raji Orat Prabakaran, BSc, Bernard Hadebe, MSc, Eric Makmur, MBBS, Guy Nicholls, MBBS; Transfusion: T Nicholas, T Parker and the Blood Transfusion laboratory team.

*Musgrove Park Hospital:* Richard Innes, MBBCh, Patricia Doble, BSc(Hons), Libby Graham, RN, Charmaine Shovelton, RN; Transfusion: M Barnett, M Davey, N Heydon, S Harlow and the Blood Transfusion laboratory team

*Nevill Hall Hospital:* Vincent Hamlyn, MBBCh, Nancy Hawkins, PhD, Anna Roynon-Reed, MSc, Sean Cutler, MSc, Sarah Lewis, MBChB; Transfusion: C Davies, J Summers, L Lewis-Prosser and the Blood Transfusion laboratory team.

*Newham University Hospital:* Juan Martin Lazaro, PhD, Tabitha Newman, MSc; Transfusion: H Mcaleese, P Winter, G Heywood-Beldon, J Lancut, C Booth, M Al-Bayati and the Blood Transfusion laboratory team.

*Ninewells Hospital:* Pauline Austin, MBChB, Susan Chapman, MBChB, Louise Cabrelli, BSc; Transfusion: K Hands, E Knight, L Macdonald.

*Norfolk and Norwich University Hospital:* Simon Fletcher, FFICM, Jurgens Nortje, FFICM, Deirdre Fottrell-Gould, Dip, Georgina Randell, Dip, Katie Stammers, BSc; Transfusion: D Asher, Janet Pring, K Ford, A Rudd and the Blood Transfusion laboratory team.

*Northampton General Hospital:* Mohsin Zaman, MRCP, Einas Elmahi, MPhil, Andrea Jones, PhD, Kathryn Hall, Dip; Transfusion: K Spreckley, M Comery, R Bisa, T Darling and the Blood Transfusion laboratory team.

*Northern General Hospital, Sheffield:* Gary H Mills, PhD, Kim Ryalls, RegPharmTech, Kate Harrington RCN, Helen Bowler, BSc, Jas Sall, BSc, Richard Bourne, PhD; Transfusion: B Taylor, S Ali, H Wilkinson, G Powell, Z Johnson and the Blood Transfusion laboratory teams.

*North Manchester General Hospital:* Zoe Borrill, MD, Tracy Duncan, MD, Thomas Lamb, MD, Joanne Shaw, BSc, Claire Fox, BSc, Kirstie Smith, BSc, Sarah Holland, Bethany Blackledge, BSc, Liam McMorrow, BSc, Laura Durrans, Jade Harris; Transfusion: T Trimble, E Cooperwaite, C Parker, S Pendlebury, J Peters, S Khan, E Anyanwu, M Evans and the Blood Transfusion laboratory teams.

*North Middlesex University Hospital:* Jeronimo Moreno Cuesta, MD, Kugan Xavier, MD, Dharam Purohit, EDIC, Munzir Elhassan, MBBS, Anne Haldeos, BSc, Rachel Vincent, DipHE, Marwa Abdelrazik, MBBCH, Samuel Jenkins, BMBS, Arunkumar Ganesan, MD, Rohit Kumar, DA, David Carter, MBBS, Dhanalakshmi Bakthavatsalam, BSc; Transfusion: S Palihavadana, K Madgwick and the Blood Transfusion laboratory team.

*Oxford University Hospitals:* Matthew Rowland, FFICM, Paula Hutton, PGCert, Archana Bashyal, MSc, Neil Davidson, BSc, Clare Hird, MSc, Sally Beer, MSc; Transfusion: J Staves, C Vander Riet, W Byrne and the Blood Transfusion laboratory team.

*Pilgrim Hospital Boston:* Manish Chhablani, FFICM, Gunjan Phalod, MPharm, Amy Kirkby, BSc(Hons), Simon Archer, BSc(Hons), Kimberley Netherton, RGN; Transfusion: B Holmes, S MacDonald, A Jackson, C Richardson and the Blood Transfusion laboratory team.

*Princess Royal Hospital:* Barbara Philips, MD, Dee Mullan, BSc, Denise Skinner, BSc, Jane Gaylard, BSc, Julie Newman, BSc; Transfusion: J Cole, L Humber and the Blood Transfusion laboratory team.

*Princess of Wales Hospital:* Sonia Arun Sathe, MD, Lisa Roche, BSc, Ellie Davies, BSc, Keri Turner; Transfusion: T Home, K Jones, R Lawton, T Edwards, C Brookes, K Hennessy, R Ward and the Blood Transfusion laboratory team.

*Poole Hospital:* Henrik Reschreiter, FFICM, Julie Camsooksai, PGDE, Sarah Patch, BSc(Hons), Sarah Jenkins, BSc(Hons), Charlotte Humphrey, BSc (Hons); Transfusion: M Greenslade, L Poole, N Dewland, M Deighton and the Blood Transfusion laboratory team.

*Queen Alexandra Hospital Portsmouth:* David Pogson, MSc, Steve Rose, BSc, Zoe Daly, BSc, Lutece Brimfield, BN, Angie Nown; Transfusion: A Davies, K Heron, G Matthias and the Blood Transfusion laboratory team.

*Queen Elizabeth Hospital, Birmingham:* Dhruv Parekh, PhD, Colin Bergin, BSc, Michelle Bates, BSc, Christopher McGhee, BSc, Daniella Lynch, BSc, Khushpreet Bhandal, Dip, Kyriaki Tsakiridou, MSc, Amy Bamford, BSc, Lauren Cooper, MSc, Tony Whitehouse, MD, Tonny Veenith, MD; Transfusion: K Wood, J Jones and the Blood Transfusion laboratory teams.

*Queen Elizabeth University Hospital, Glasgow:* Malcolm A.B. Sim, MD, Sophie Kennedy Hay, BN, Steven Henderson, MPH, Maria Nygren, MSc, Eliza Valentine, HNC; Transfusion: C McKie, A Hanlon, A Molloy, M McGarvey and the Blood Transfusion laboratory team.

*Queen's Hospital, Burton:* Amro Katary, MD, Gill Bell, BSc, Louise Wilcox, BSc, Katy English BSc, Ann Adams; Transfusion: J Buchan, J Jeyachandran, R Shiers, S Kaur, H Ahmad, H Clarke, K Kacinova and the Blood Transfusion laboratory teams.

*Queen's Hospital, Romford:* Mandeep-Kaur Phull, MBBS, Abbas Zaidi, MBBS, Tatiana Pogreban, BN, Lace Paulyn Rosaroso, BN; Transfusion: X Tang, A Minogue, A Abdulle, N Munu, N Page, S Balkee, P Patel, C Sanderson, R Gad, A Asquith, A Minhas, G Marange, S Lockyer

*Queens Medical Centre and Nottingham City Hospital:* Daniel Harvey, BMBS, Benjamin Lowe, BMBS, Megan Meredith, BSc(Hons), Lucy Ryan, MNSc, DREEAM Research Team; Transfusion: M Skill, N Horton-Turner, A Tervit, C Wilkes, R Rose, S Springworth, H Bond, S Robinson, L Allen, C Chang, M I Saez-Garcia-Holloway and Blood Transfusion laboratory team.

*The Rotherham NHS Foundation Trust:* Anil Hormis, FRCA, Rachel Walker, BA, Dawn Collier, BSc, Sarah Kimpton, MSc, Susan Oakley; Transfusion: R Stirk, R Perkins, S Lord, C Bilton and the Blood Transfusion laboratory team.

*Royal Alexandra Hospital:* Kevin Rooney, MBChB, Natalie Rodden, BSc, Emma Hughes, BSc, Nicola Thomson, BSc(Hons), Deborah McGlynn, BSc, Charlotte Clark, Dip, Patricia Clark, BSc; Transfusion: Hospital Blood Transfusion Team

*Royal Berkshire Hospital:* Andrew Walden, FFICM, Liza Keating, MBChB, Matthew Frise, DPhil, Tolu Okeke, BSc, Nicola Jacques, MSc, Holly Coles, BSc, Emma Tilney, BSc, Emma Vowell, DipHE; Transfusion: K Kemsley, T Hawkins and the Blood Transfusion laboratory team.

*Royal Bournemouth and Christchurch Hospitals:* Martin Schuster-Bruce, FRCA, Sally Pitts, BSc, Rebecca Miln, ADipHE, Laura Purandare, MBA, Luke Vamplew, BSc; Transfusion: M Trevett, A Jose, L Mounsey, C Baylem, T Haydon, V Chandler-Vizard, B Grice and the Blood Transfusion laboratory team.

*Royal Brompton Hospital:* Brijesh Patel, FRCA, Debra Dempster, Mahitha Gummadi, Natalie Dormand, Shu Fang Wang; Transfusion: V Jeyakumar, H Day, C Biring, P Agent, M Gaspar and the Blood Transfusion Laboratory teams

*Royal Cornwall NHS Trust:* Michael Spivey, FFICM, Sarah Bean, RN, Karen Burt, RN, Lorraine Moore, MPharm; Transfusion: A Parsons and the Blood Transfusion laboratory team.

*Royal Devon and Exeter NHS Foundation Trust:* Christopher Day, MD, Charly Gibson, MBChB, Elizabeth Gordon, BSc, Letizia Zitter, BSc, Samantha Keenan, BSc; Transfusion: J Davies, J Piper and the Blood Transfusion laboratory team.

*Royal Glamorgan Hospital:* Jayaprakash Singh, MD, Ceri Lynch, MD, Lisa Roche, Justyna Mikusek, Bethan Deacon, Keri Turner; Transfusion: T Home, K Jones, R Lawton, T Edwards, C Brookes, K Hennessy, R Ward and the Blood Transfusion laboratory team.

*Royal Gwent Hospital*: Tamas Szakmany, PhD, Evelyn Baker, MSc, Shiney Cherian, BSc(Hons), John Hickey, MSc, Shreekant Champanerkar, MBBS ; Transfusion: C Davies, J Summers, L Lewis-Prosser and the Blood Transfusion laboratory team.

*Royal Hallamshire Hospital, Sheffield:* Gary H Mills, PhD, Ajay Raithatha, FFICM, Kris Bauchmuller, FFICM, Norfaizan Ahmad, FFICM, Matt Wiles FFICM, Jayne Willson, RN; Transfusion: B Taylor, S Ali, H Wilkinson, G Powell, Z Johnson and the Blood Transfusion laboratory teams.

*Royal Hampshire Hospitals:* Irina Grecu, MD, Jane Martin, Caroline Wrey Brown, Ana-Marie Arias, Emily Bevan; Transfusion: M Cundall, O Martins, C Wilson, L Holloway and the Blood Transfusion laboratory team.

*Royal Infirmary of Edinburgh:* Thomas H Craven, PhD, David Hope, PGDip, Jo Singleton, BN, Sarah Clark, MNurs, Corrienne McCulloch, PhD; Transfusion: J Falconer, J Oldham and the Blood Transfusion laboratory team.

*Royal Liverpool University Hospital:* Ingeborg D Welters, PhD, David Oliver Hamilton, BMBS, Karen Williams, RGN, Victoria Waugh, BA, David Shaw, DipHE, Suleman Mulla, MBChB, Alicia Waite, PhD, Jaime Fernandez Roman, BSc, Maria Lopez Martinez, BSc; Transfusion: K Knowles, H Wissett, R Wright, C Dragomir, A Peacock, J Bell, J Gorry, I Houghton, P Burgess and all members of the lab BMS and MLA teams.

*Royal London Hospital:* Zudin Puthucheary, PhD, Timothy Martin, BA(Hons), Filipa Santos, RN, Ruzena Uddin, MSc(Hons), Maria Fernandez, MSc, Fatima Seidu, MSc, Alastair Somerville, MSc, Mari Lis Pakats, MSc, Priya Dias, PhD, Salam Begum, BSc, Tasnin Shahid, BSc; Transfusion: H Mcaleese, P Winter, G Heywood-Beldon, J Lancut, C Booth, M Al-Bayati and the Blood Transfusion laboratory team.

*The Royal Free Hospital:* Sanjay Bhagani, FRCP, Mark De Neef, MSc, Helder Filipe, BSc, Sara Mingos, BSc, Amitaa Maharajh, BA, Glykeria Pakou, BA, Aarti Nandani, MPharm; Transfusion: R Atugonza, A Li, J Li, S Tekle, A M Ellis, A Natarajan, S Mellin and the Blood Transfusion laboratory team.

*The Royal Marsden NHS Foundation Trust:* Kate Colette Tatham, PhD, Shaman Jhanji, PhD, Ethel Black, BSNurs, Arnold Dela Rosa, BSNurs, Ryan Howle, FRCA, Ravishankar Rao Baikady, FRCA; Transfusion: E Malundas, A Mohamed, L Desai.

*The Royal Oldham Hospital:* Redmond P Tully, FFICM, Andrew Drummond, FFICM, Joy Dearden, BSc, Jennifer E Philbin, MSc, Sheila Munt, SRN; Transfusion: A Allameddine, J Uttley, S Andrews, C Porada, K Mushtaq, D Curley, H Morris, O Akinwumiju, C Okyne-Turkson, S Flynn and the Blood Transfusion laboratory teams.

*The Royal Wolverhampton NHS Trust:* Shameer Gopal, MBBCh, Jagtar- Singh Pooni, MBBS, Saibal Ganguly, MBBS, Andrew Smallwood, RGN, Stella Metherell, RGN; Transfusion: M Boyd, M Blanton, M Herbert and the Blood Transfusion Laboratory team.

*Royal Papworth Hospital:* Alain Vuylsteke, MD, Charles Chan, FRCA, Saji Victor, COVID Research Team, Papworth Hospital; Transfusion: M Muir, J Joseph, C Flatters, A Hudson and the Biomedical Scientists and Associate Practitioners of the Transfusion laboratory.

*Royal Stoke Hospital:* Ramprasad Matsa, FRCP, Minerva Gellamucho, BSN, Michelle Davies, NVQ; Transfusion: R Sivers, J Graham, P Irving, D Bentley, A Salmon, R Rushworth, C Brackstone, C Baker, S Mitchell, E Brown, S O'Brien, J Jeffrey, F Perkins, D Murdoch and the Blood Transfusion laboratory teams.

*Royal Surrey County Hospital:* Ben Creagh-Brown, PhD, Joe Tooley, MSc, Laura Montague, BSc, Fiona De Beaux, BSc, Laetitia Bullman, MBChB; Transfusion: S Vimalanathan, J Lawrence and the Blood Transfusion laboratory team.

*Royal United Hospital Bath:* Ian Kerslake, FFICM, Carrie Demetriou, RN, Sarah Mitchard, MBBS, Lidia Ramos, RN, Katie White, MSc; Transfusion: A Wardle, A Dornan, W Vietri, K Pass, H Maria-Osborn, H Ali, C Risbridger, A Buchanan, K Turek, D Loveys, L Swan, R Coles, P Perera, Z Sibanda and the laboratory team.

*Russells Hall Hospital:* Michael Reay, FFICM, Steve Jenkins, MD, Caroline Tuckwell, Angela Watts, BSc, Eleanor Traverse, Stacey Jennings; Transfusion: A Smith, C Tuckwell and the Blood Transfusion laboratory team.

*Salisbury NHS Foundation Trust:* Phil Donnison, FFICM, Maggie Johns, RGN, Ruth Casey, BSc, Lehentha Mattocks, Dip, Sarah Salisbury; Transfusion: C Matthews, S Haviland, C Stacey and the Blood Transfusion laboratory team.

*Salford Royal NHS Foundation Trust:* Paul Dark, PhD, Alice Harvey, BSc, Reece, Doonan, BSc, Liam McMorrow, BA (Hons), Karen Knowles, BA (Hons);*Sandwell and West Birmingham NHS Trust:* Jonathan Hulme, FFICM, Santhana Kannan, FFICM, Sibet Joseph, BSc, Fiona Kinney, RGN, Ho Jan Senya, BPharm; Transfusion: D Seddon, J Wesson, L Baxter, L Cooper, E Loutraris, all Haematology and Blood Transfusion staff.

*Sherwood Forest Hospitals NHS Foundation Trust:* Valli Ratnam, MD, Mandy Gill, Jill Kirk, Sarah Shelton; Transfusion: J Wren, J Walden, L Hostler, Senior BMS and the Blood Transfusion laboratory team.

*South Tyneside District Hospital:* Christian Frey, MD, Riccardo Scano, MD, Madeleine McKee, BSc, Peter Murphy, BSc; Transfusion: L Sudlow, J Caulfield, J Trattles, P Patterson, S Matthews and all in the Blood Transfusion laboratory teams.

*Southmead Hospital:* Matt Thomas, FFICM, Ruth Worner, RGN, Beverley Faulkner, RGN, Emma Gendall, BSc, Kati Hayes, BSc, Hayley Blakemore, BSc, Borislava Borislavova, MSc; Transfusion: T Wreford-Bush, K Mead, A Morley and the Blood Transfusion laboratory team.

*St. Bartholomew's Hospital:* Colin Hamilton-Davies, MBBS, Carmen Chan, BSc, Celina Mfuko, BSc, Hakam Abbass, MSc, Vineela Mandadapu, MSc; Transfusion: H Mcaleese, P Winter, G Heywood-Beldon, J Lancut, C Booth, M Al-Bayati and the Blood Transfusion laboratory team.

*St. George's Hospital:* Susannah Leaver, MRCP, Kamal Patel, MRCP, Sarah Farnell-Ward, MSc, Romina Pepermans Saluzzio, BSc, John Rawlins, MBBS; Transfusion: C Orchard, V Michael, K Feane, J Uprichard and all the transfusion lab team.

*St. Mary's Hospital:* Anthony Gordon, MD; Dorota Banach, BSc, Ziortza Fernández de Pinedo Artaraz, BN, Leilani Cabreros, BSN; Transfusion: A Rahman, D Johnson, L Chapple, E Nweje, K Sabljak, S Kassa, U Munu, H Dawson, F Regan, F Chowdhury, V Chiroma and Blood Transfusion laboratory teams.

*St. Peter's Hospital, Chertsey:* Ian White, FFICM, Maria Croft, BSc(Hons), Nicky Holland, BN(Hons), Rita Pereira, MPharm; Transfusion: J Finden, Z Takats, N Johnson and the Blood Transfusion Laboratory team.

*Stepping Hill Hospital, Stockport:* Ahmed Zaki, PhD, David Johnson, MPhil, Matthew Jackson, MBChB, Hywel Garrard, BMBS, Vera Juhaz, MD, Louise Brown BSc (Hons); Transfusion: R Zaman, B Devine and the Blood Transfusion laboratory team.

*Sunderland Royal Hospital:* Alistair Roy, MBChB, Anthony Rostron, PhD, Lindsey Woods, BSc, Sarah Cornell, BSc; Transfusion: L Sudlow, J Caulfield, J Trattles, P Patterson, S Matthews and all in the Blood Transfusion laboratory teams.

*Swansea Bay University Health Board:* Suresh Pillai, FFCIM, Rachel Harford, RN, Helen Ivatt, FRCA, Debra Evans, BN, Suzanne Richards, BN, Eilir Roberts, MBBCh, James Bowen, MBBCh James Ainsworth, MBBS; Transfusion: D Payne, K Phillips, L Park, S John, P Diamond and the Blood Transfusion laboratory team.

*Torbay and South Devon NHS Foundation Trust:* Thomas Clark, MBChB, FRCA, FFICM, Angela Foulds, BSc, Stacey Atkins, Dip RN; Transfusion: A Penny, S Mills, J Pinder, H Randall, M Fisher Crisp and the Blood Transfusion laboratory team.

*United Lincolnshire NHS Trust:* Kelvin Lee, PhD, Russell Barber, FRCA, Anette Hilldrith, RGN, Claire Hewitt, RGN, Gunjan Phalod, MPharm; Transfusion: B Holmes, S MacDonald, A Jackson, C Richardson and the Blood Transfusion laboratory team.

*University Hospitals Coventry & Warwickshire NHS Trust:* Pamela Bremmer, BSc, Geraldine Ward, MA, Christopher Bassford, PhD; Transfusion: A Busby, J Northcote and the Blood Transfusion laboratory team.

*University Hospital of North Tees:* Farooq Brohi, FFARCSI, Vijay Jagannathan, FRCA, Michele Clark, MA, Sarah Purvis, Dip, Bill Wetherill, MSc; Transfusion: M T Walker, D A Cox and the Blood Transfusion laboratory team.

*University Hospital Southampton NHS Foundation Trust:* Ahilanandan Dushianthan, PhD, Rebecca Cusack, MD, Kim de Courcy-Golder, PGDip, Karen Salmon, MSc, Rachel Burnish, Simon Smith, BN, Susan Jackson, BSc, Winningtom Ruiz, BSc (Hons), Zoe Duke BSc (Hons) Magaret Johns, BA, Michelle Male, Dip, Kirsty Gladas, BSc (Hons), Satwinder Virdee, MPharm, Jacqueline Swabe, MPharm, Helen Tomlinson; Transfusion: K Dowling, J Ricks, S Carrington, P Downey, S Mumford, T Lofting and the Blood Transfusion laboratory team.

*Warwick Hospital:* Ben Attwood, MBBCh, Penny Parsons, BSc, Bridget Campbell, BSc, Alex Smith, BSc;

*Watford General Hospital:* Valerie J Page, MBBCh, Xiao Bei Zhao, BSc (Hons), Deepali Oza, BPharm, Gail Abrahamson, DPhil, Ben Sheath, BSc (Hons), Chiara Ellis, BSc (Hons); Transfusion: D Beckford-Smith, S Bradley

*Western General Hospital, Edinburgh:* Jonathan Rhodes, PhD, Thomas Anderson, MBChB, Sheila Morris; Transfusion: J Falconer, J Oldham and the Blood Transfusion laboratory team.

*Whipps Cross Hospital:* Charlotte Xia Le Tai, MBChB, Amy Thomas, MSc, Alexandra Keen, MSc; Transfusion: H Mcaleese, P Winter, G Heywood-Beldon, J Lancut, C Booth, M Al-Bayati and the Blood Transfusion laboratory team.

*Whiston Hospital*: Dr Ascanio, Tridente, Karen Shuker, Jeanette Anders, Sandra Greer, Paula Scott, Amy Millington, Philip Buchanan, Jodie Kirk; Transfusion: M Wood, T Kelly, S Adair, V Hayes and the Blood Transfusion laboratory team.

*Wirral University Teaching Hospital NHSFT:* Craig Denmade, MBChB, Girendra Sadera, MBBS, Reni Jacob, BSc, Cathy Jones, BSc, Debbie Hughes; Transfusion: S Carter, L Delaney, D Woods and the Blood Transfusion laboratory team.

*Worcester Royal Hospital:* Stephen Digby, MBBS, Nicholas Cowley, MD, Laura Wild, BSc(Hons), Jessica Thrush, RGN, Hannah Wood, BSc(Hons), Karen Austin, RGN; Transfusion: C Khan, G Godding, E Murphy, E Loxley and the Blood Transfusion laboratory team.

*Wrexham Maelor Betsi Cadwaladr University Hospital*: David Southern, FFICM, Harsha Reddy, FFICM, Sarah Hulse, BSc, Andy Campbell, FFICM, Mark Garton, Claire Watkins, PGDip, Sara Smuts, BN; Transfusion: T Coates, S Thomas-Wright, C Bennett and the Blood Transfusion laboratory team

*Wrightington, Wigan and Leigh Teaching Hospitals NHS Foundation Trust:* Alison Quinn, MD, Benjamin Simpson, MD, Catherine McMillan, MD, Cheryl Finch, BSc, Claire Hill BSc, Josh Cooper; Transfusion: L McCreery, J Cooper, J Wesson and the Blood Transfusion laboratory team.

*Wye Valley NHS Trust:* Joanna Budd, MBBS, Charlotte Small, PhD, Ryan O'Leary, MBBS, Janine Birch, RN, Emma Collins, BSc (Hons); Transfusion: I Hancock, H Slade, M Kevern and the Blood Transfusion laboratory team.

*Wythenshawe Hospital*: Peter D G Alexander, FFICM, Tim Felton, FFICM, Susan Ferguson, BSc, Katharine Sellers, BSc, Joanne Bradley-Potts, BSc; Transfusion: T Trimble, E Cooperwaite,

C Parker, S Pendlebury, J Peters, S Khan, E Anyanwu, M Evans and the Blood Transfusion laboratory teams.

*York Teaching Hospital*: David Yates, FCRA, Isobel Birkinshaw, BSc(Hons), Kay Kell, BSc(Hons), Zoe Scott, BN, Harriet Pearson, BSc; Transfusion: C Purcell, A Thorpe, C Ivel, J Fullthorpe and the Blood Transfusion laboratory team.
